# His-MMDM: Multi-domain and Multi-omics Translation of Histopathological Images with Diffusion Models

**DOI:** 10.1101/2024.07.11.24310294

**Authors:** Zhongxiao Li, Tianqi Su, Bin Zhang, Wenkai Han, Sibin Zhang, Guiyin Sun, Yuwei Cong, Xin Chen, Jiping Qi, Yujie Wang, Shiguang Zhao, Hongxue Meng, Peng Liang, Xin Gao

**Affiliations:** Computer Science Program, Computer, Electrical and Mathematical Sciences and Engineering Division, King Abdullah University of Science and Technology (KAUST), Thuwal, Saudi Arabia; Center of Excellence on Smart Health, King Abdullah University of Science and Technology (KAUST), Thuwal, Saudi Arabia; Department of Neurosurgery, Harbin Medical University Cancer Hospital, Harbin, China; Department of Pathology, The First Affiliated Hospital of Harbin Medical University, Harbin, China; Department of Neurosurgery, The First Affiliated Hospital of Harbin Medical University, Harbin, China; Department of Pathology, Harbin Medical University Cancer Hospital, Harbin, China

**Keywords:** Image Generative Models, Image Translation, Diffusion Models, Histopathology

## Abstract

Generative AI (GenAI) has advanced computational pathology through various image translation models. These models synthesize histopathological images from existing ones, facilitating tasks such as color normalization and virtual staining. Current models, while effective, are mostly dedicated to specific source-target domain pairs and lack scalability for multi-domain translations. Here we introduce His-MMDM, a diffusion model-based framework enabling multi-domain and multi-omics histopathological image translation. His-MMDM is not only effective in performing existing tasks such as transforming cryosectioned images to FFPE ones and virtual immunohistochemical (IHC) staining but can also facilitate knowledge transfer between different tumor types and between primary and metastatic tumors. Additionally, it performs genomics-and/or transcriptomics-guided editing of histopathological images, illustrating the impact of driver mutations and oncogenic pathway alterations on tissue histopathology and educating pathologists to recognize them. These versatile capabilities position His-MMDM as a versatile tool in the GenAI toolkit for future pathologists.

## Introduction

Generative artificial intelligence (GenAI) in imagery has marked a groundbreaking advance in the field of artificial intelligence (AI), offering transformative capabilities across a wide range of applications. At the forefront, the state-of-the-art text-to-image models such as Stable Diffusion ^1^, Midjourney ^2^, and DALL-E 3 ^3^, transform text descriptions into vivid visual representations. Conversely, image-to-text models, such as GPT-4V ^4^, interpret and convert visual data into descriptive text, enabling open-ended dialogs about the image content with users. There are also image-to-image translation models that use existing images as templates to generate new ones, enabling applications such as image editing, resolution enhancement, and sketch-to-image synthesis ^1,5^. Collectively, these models are revolutionizing the way we create, interpret, and interact with visual contents.

Such a revolution is also gradually taking place in the field of computational pathology. Early explorations have demonstrated the effectiveness of using generative adversarial networks (GANs) ^6^ or diffusion models (DMs) ^7^ to generate synthetic histopathological images ^8–10^, and have been mainly used as a data augmentation strategy ^11–13^. Recently, a lot of efforts have been devoted to improving the quality and enlarging the resolution of the generated images ^14,15^. Concurrently, visual language foundation models and copilot systems are also being developed in computational pathology to automate medical visual question answering ^16–20^. As for image-to-image translation, dedicated models have been developed for various common tasks such as color normalization ^21^, stain transfer ^22^, virtual staining ^22–25^, and transforming cryo-sectioned or stimulated Raman images to formalin-fixed paraffin-embedded (FFPE) ones ^26,27^.

In general, there are two types of image-to-image translation tasks, including the paired and unpaired ones. The paired image translation task requires matched source and target domain images during training, which is usually not readily available ^28^. In contrast, the unpaired image translation task requires the algorithm to learn the mapping between a source and target domain on itself, making it more useful in real-world applications ^28^. It is thus not surprising that almost all the existing image translation models in computational pathology are un-paired. Although the abovementioned image translation models serve as important computational pathology applications, their full potential still has not yet been unleashed. Firstly, current applications are mostly limited to GANs, whereas recent developments of DMs show their superior performance on conditional generation as compared to GANs ^29^. Secondly, the current GAN-based histopathological image translation models are limited to the translation between a couple of specific source-target domain pairs and their extension to the translation between multiple domains is not straightforward ^30^. Hence, simultaneous translation across multiple domains has a wider range of applications in computational pathology. For example, one can edit one real histopathological image into multiple synthesized versions corresponding to altered genomic or transcriptomic profiles, which can potentially model the impact of mutations and altered gene expression on the histopathological appearance of the tissue. Furthermore, multiple-domain translations may also alleviate the inefficiency of pairwise translation, reducing the number of *n*(*n* − 1)/2 individual pairwise models to one multi-domain model. Thirdly, current GAN-based models have specifically designed components or loss functions that may hinder their extensibility to new applications ^25,26^.

Due to the above limitations, in this study, we propose a novel versatile framework, Histopathological image Multi-domain Multi-omics translation with Diffusion Models (His-MMDM). His-MMDM achieves image translation by diffusing the source domain images into noisy images distributed as Gaussian distribution and then denoising the noisy images back into target domain images. Compared with previous image translation models in histopathology, His-MMDM stands out with two unique capabilities: (1) it can be efficiently trained to translate images between an unlimited number of categorial domains (through class-conditional inputs to the model) (2) it can perform genomics-or transcriptomics-guided editing of histopathological images (through multi-omics inputs to the model). For categorical domain translation, we demonstrated His-MMDM’s performance through four comprehensive tasks using four independent datasets, including the translation of cryo-sectioned slide images to FFPE ones, virtual immunohistochemical (IHC) staining of hematoxylin and eosin (H&E) images, primary tumor type translation, and tumor organ site translation. Most notably, we showed through comprehensive experiments that His-MMDM’s generated contents can improve the performance of recent histopathological foundation models ^31–33^ that were mainly trained on FFPE images on various discriminative and retrieval tasks. It can also facilitate histopathological knowledge transfer between different tumor types and primary and metastatic tumors. Using the histopathological images in The Cancer Genome Atlas (TCGA) that are equipped with the matched genomic and transcriptomic profiles, we demonstrated His-MMDM’s effectiveness in performing genomics– and transcriptomics-guided editing of the images. We then systematically discussed and illustrated through notable examples the effect of high-frequency somatic mutations and alterations on the expression level of genes/pathways on the histopathology of the tissue from a generative model’s perspective. We then demonstrated the usefulness of these AI-generated contents (AIGC) by showing that they are useful in educating pathologists to recognize these underlying conditions. Overall, we believe that His-MMDM showcases the possibility of generic multi-domain and multi-omic translation and generation in histopathology, provides a powerful tool for pathologists, and could serve as an integral component of GenAI copilot systems in the future.

## Results

### The design of His-MMDM model

His-MMDM is trained based on the principle of diffusion models (DMs) ^7^ and is trained with histopathological images. A DM is comprised of a forward diffusion and a backward denoising process (**Fig. 1**). In the forward diffusion procedure, a histopathological image (*X*_*T*_) is gradually diffused into a noisy image (*X*_0_) following a Gaussian distribution using a pre-determined schedule and in the backward denoising procedure, the noisy image is gradually denoised back into a histopathological image (**Methods**). At each iteration of the denoising procedure, a U-net-based ^7,34^ network is used to predict noise from the image at the current iteration step, which is utilized in the subsequent denoising step derived from the inversion of the forward procedure (**Methods**) ^35^. Conditional generation is achieved by the addition of condition information to the U-net through embedding vectors added to each layer of the U-net blocks (**Fig. 1**, **Table S1**, **Methods**). The DM used in His-MMDM can handle three types of condition information: (1) categorical labels, such as FFPE/cryosection, IHC stains, tumor types, and tumor primary sites. (2) genomics, which includes the mutation status of the corresponding sample, and (3) transcriptomics, which includes the transcriptomic profile of the sample. After His-MMDM is trained, it achieves image translation by (1) executing the forward diffusion procedure of the trained DM by conditioning the U-net with the source domain condition information and then (2) executing the backward denoising procedure by conditioning the U-net with the target domain condition information (detailed in **Methods**). We demonstrate in the following sections that such a strategy can successfully achieve multi-domain and multi-omics image translation.

**Figure 1.**
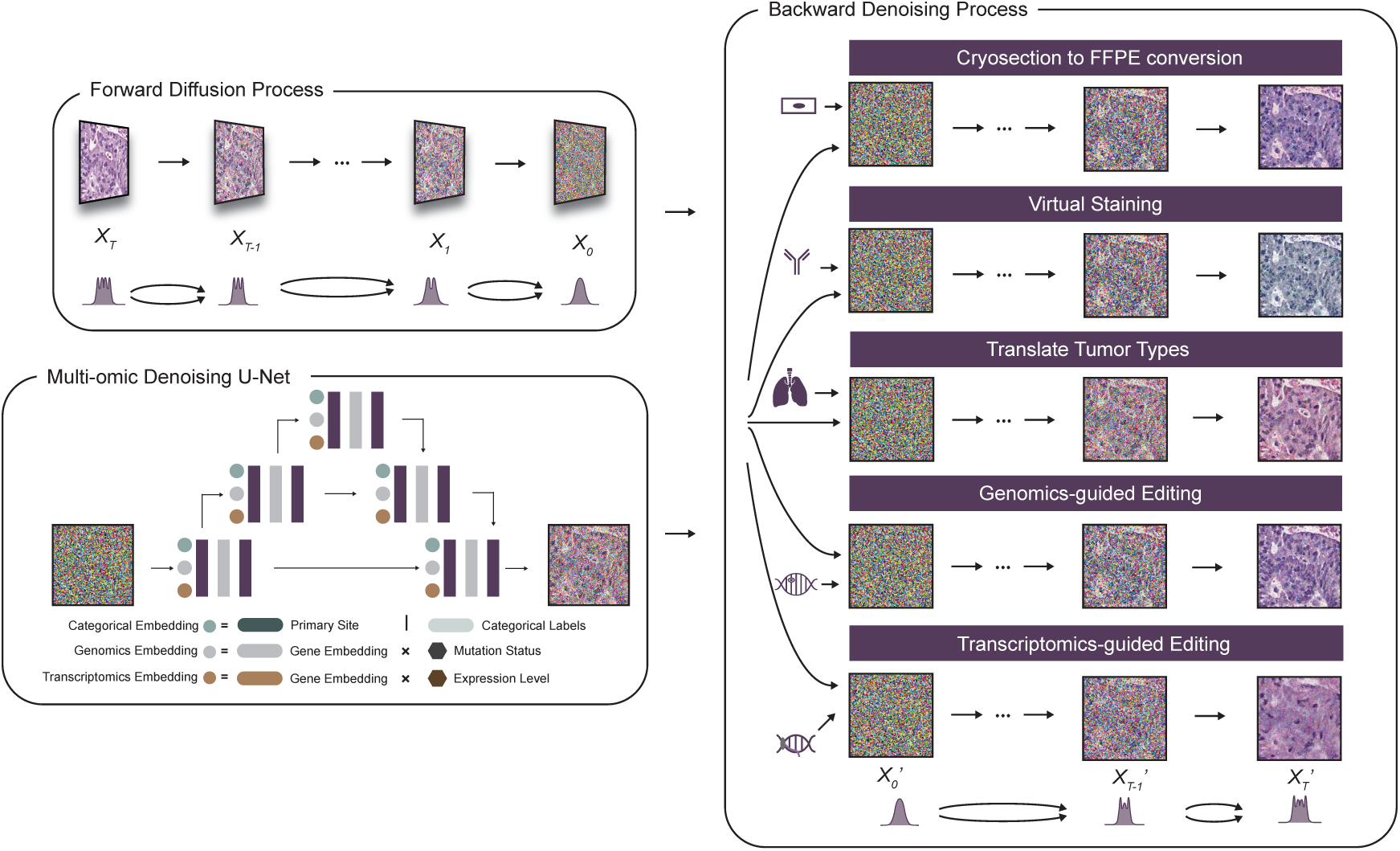
Architecture of His-MMDM. The architecture of Histopathological image Multi-domain Multi-omic translation with Diffusion Models (His-MMDM). Based on the principle of DMs, the forward diffusion procedure gradually diffuses a histopathological image into a noisy image following a Gaussian distribution. The backward procedure takes in the noisy image and gradually denoises it back into a real histopathological image. A U-net-based network makes noise prediction for a given input and takes in condition information in the format of categorical labels, genomic profiles, and transcriptomic profiles in its embedding layers.

### Cryosection to FFPE conversion and virtual IHC staining of histopathological images

We first evaluated the effectiveness of His-MMDM on previously studied image translation tasks, such as converting cryosectioned slide images to formalin-fixed and paraffin-embedded (FFPE) images ^26^, as well as virtual IHC staining ^24,25,36^, without any task-specific adaptation of the model architecture.

To convert cryosectioned slide images into FFPE with His-MMDM, we trained it to conditionally generate TCGA slide images of their respective slide type. We collect histopathological image patches from a total of 19 tumor types from five families, including five gastrointestinal (GI) (ESCA, STAD, COAD, READ, and PAAD), two lung (LUAD and LUSC), three kidney (KIRP, KICH, and KIRC), three pan-gynecological (GYN) (OV, BRCA, and UCEC), and five other tumor types (LIHC, HNSC, SARC, THCA, BLCA, and PRAD) (**Methods**, statistics shown in **Table S2**, abbreviations defined in **Table S3**). We ran the forward diffusion process on a cryosectioned slide image under the condition ‘cryosection’ followed by the backward denoising process under the condition ‘FFPE’ (**Fig. 2a, Methods, Table S1**). We translated the test image patches from all 19 tumor types in TCGA using our pre-trained model and evaluate the reduction of the Frechet Inception Distance (FID) before and after translation (ΔFID = FID_before_ − FID_after_) as the metric for evaluating the performance of translation (**Fig. 2b**). Although His-MMDM is not specifically designed for this task, it achieved performance comparable to the dedicated method AI-FFPE ^26^ across most tumor types, even surpassing it in six out of 19 tumor types. In the remaining tumor types where AI-FFPE has an edge, the performance gap between His-MMDM and AI-FFPE is not greater than that observed with other methods. Overall, His-MMDM achieved better performance compared to other general GAN-based models, CycleGAN ^28^ and CUT ^37^. For other diffusion model-based image translation methods, His-MMDM achieved comparable performance as BBDM ^38^, but is much better than the few-shot translation model D2C ^39^. Through more detailed visual examination, we found that His-MMDM can alleviate the problem of tissue dehydration and nuclear deformation that are common in cryosectioned slides and generate a better presentation of contrasting regions and arrangement of the histopathological contents (**Fig. 2c**).

**Figure 2.**
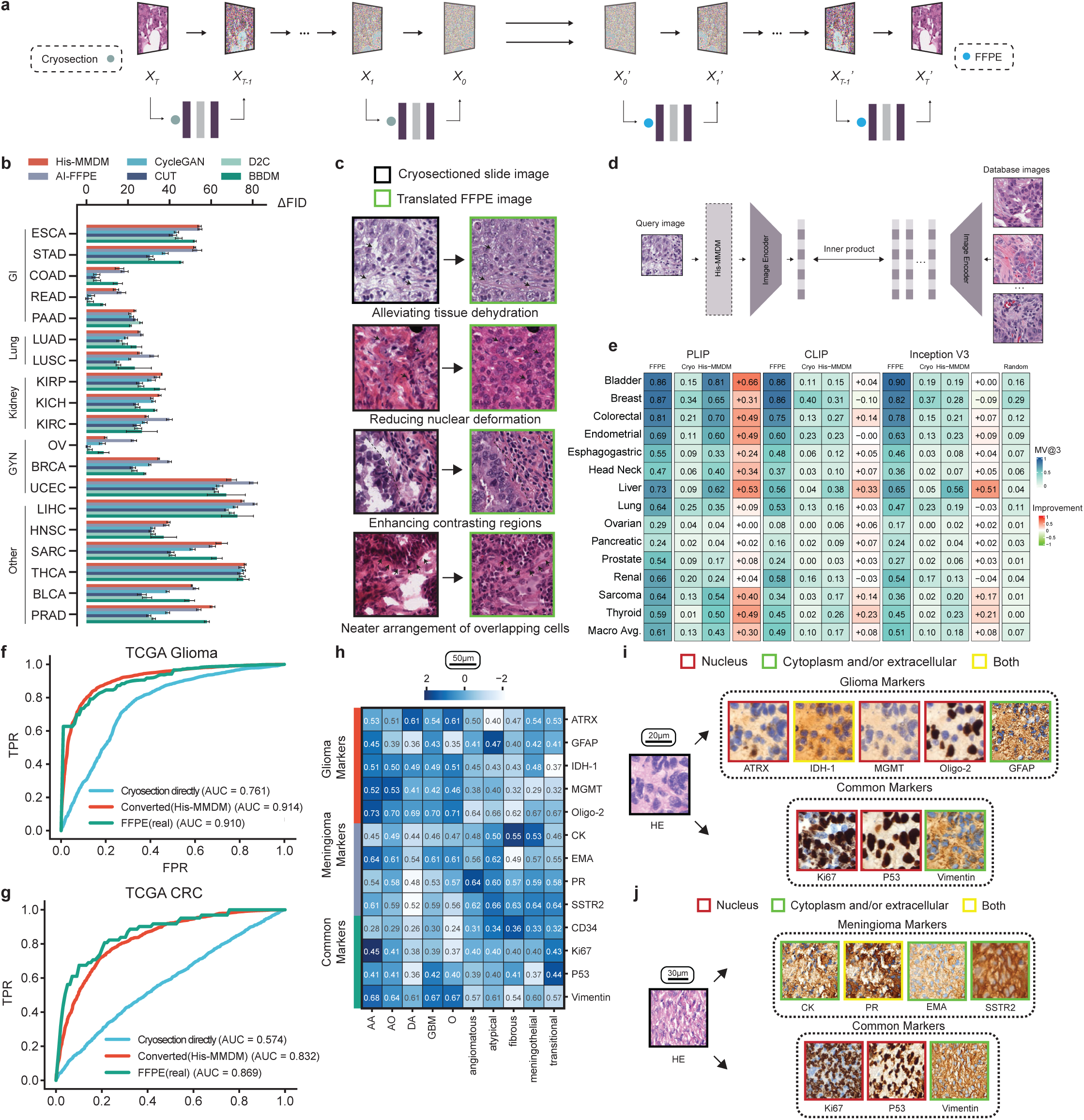
Cryosection to FFPE conversion and virtual IHC staining of histopathological images. (a) A schematic that illustrates how His-MMDM achieves cryosection to FFPE conversion. (b) Evaluation of His-MMDM (in terms of ΔFID scores) in performing cryosection to FFPE conversion and comparison with other dedicated (AI-FFPE) and non-dedicated GAN-based (CycleGAN and CUT) and diffusion model-based (D2C and BBDM) image translation models. (c) Examples of cryosection to FFPE conversion. (d) The schematic illustrating the usage of His-MMDM converted cryosectioned slide images for image-to-image retrieval using pre-trained histopathological foundation models. (e) His-MMDM converted cryosectioned slide images improve the image retrieval performance (in terms of MV@3) of the pre-trained foundation models, PLIP, CLIP, and Inception V3. FFPE: using FFPE images, Cryo: using original cryosection images, His-MMDM: using His-MMDM translated images (f-g) His-MMDM converted cryosectioned slide images improve the performance of the foundation model, CHIEF, on IDH1 mutation prediction in glioma (f) and MSI prediction in CRC (g). (h) The ‘inverted normalized FID scores’ of the virtually stained images for each IHC marker in each glioma and meningioma subtype. (i-j) Examples of virtual staining of H&E histopathological images in glioblastoma (i) and atypical meningioma (j). His-MMDM can perform virtual staining of the glioma-specific markers, the meningioma-specific markers, as well as the common markers used by both primary brain tumor types. The positive regions (indicated by the brown-colored DAB stain) are also in agreement with the respective markers’ expected location of expression (nucleus, cytoplasm, and extracellular matrix).

We emphasize that translating cryosectioned slide images into FFPE images has broader implications beyond merely improving image quality for pathologists’ observation. Although recent histopathological foundation models ^16,32,33^ are pre-trained on diverse multi-center cohorts, the majority of their training data consists of FFPE slide images rather than cryosectioned ones. Consequently, directly applying these models to cryosectioned slide images risks significant performance degradation. We systematically assessed whether this degradation could be mitigated by first translating cryosectioned slides into FFPE images using His-MMDM. Following a similar image-to-image retrieval method as in the PLIP paper ^16^, we evaluated the performance of various image encoders on an image-to-image retrieval task (**Fig. 2d**, **Methods**). With His-MMDM, the retrieval accuracy increased by 0.30, 0.08, and 0.08 for pretrained PLIP, CLIP ^40^, and InceptionV3 ^41^, respectively (**Fig. 2e**). Additionally, His-MMDM improved the text-guided classification accuracy of PLIP by 0.39 (macro-averaged, **Methods**, **Fig. S1a-b**). Beyond multi-modal pre-trained models like PLIP, His-MMDM also enhances self-supervised image-only pre-trained models, such as CHIEF ^33^ (**Methods**), on tasks such as inferring microsatellite instability (MSI) in colorectal (CRC) tumors (**Fig. 2f**) and IDH1 mutation status in gliomas (**Fig. 2g**).

For IHC virtual staining of histopathological images, we collected patch images of 13 common markers used in brain tumor diagnosis spanning five glioma subtypes and five meningioma subtypes from The First Affiliated Hospital of Harbin Medical University (hereafter referred to as the ‘HMU-1st dataset’) (**Methods**, **Table S2**). Of the 13 IHC markers, five are routinely used in glioma, four in meningioma, and four for both two (**Table S4**). In this dataset, the H&E slides and the IHC slides of each patient are simultaneously available. His-MMDM is then trained on this dataset to perform conditional generation of both H&E images as well as each of the 13 IHC markers. It achieves virtual staining by running in serial the forward diffusion process conditioned on the label ‘H&E’ and the backward denoising process conditioned on the label of the desired IHC marker (**Fig. S1c**).

We evaluated the ‘inverted normalized FID scores’ (**Methods**) of the translated images in each subtype of glioma and meningioma (**Fig. 2h**). The IHC markers into which the H&E images are translated tend to have higher performance when the marker is more compatible with the brain tumor type/subtype. For instance, the glioma markers (GFAP, ATRX, IDH-1, MGMT, and Oligo-2) tend to have higher performance in gliomas and so do the meningioma markers (EMA, PR, and SSTR2) in meningiomas (**Fig. 2h**). The positive regions of the virtually stained images (as indicated by the brown-colored diaminobenzidine (DAB) stain) are both in agreement with the marker’s expected location of expression (in the nucleus or the cytoplasm and extracellular matrix) (**Fig. 2i**-**2j, Fig. S1d-S1e**), as well as the corresponding glioma subtype (**Fig. S1f-S1h**, **Section S1**).

### Knowledge transfer of histopathological images across primary tumor types

We then experimented with His-MMDM to translate histopathological images of one tumor type to another. The categorical domains for this task are the above-mentioned 19 primary tumor types of The Cancer Genome Atlas (TCGA) (**Methods**). Using discriminative models, previous studies ^42,43^ demonstrated high cross-classification performance on a lot of these tumor type pairs, i.e., the performance of a tumor classifier trained on one tumor type and tested on another, which suggests conserved tumor image features across different types of tumors.

We tasked His-MMDM to translate the histopathology images between each two across these 19 primary tumor types (**Fig. 3a**). We systematically evaluated the translation performance of His-MMDM and other baseline methods and discussed the heterogenous performance across different translation pairs (**Fig. S2a-S2b, Section S2**). Through visual inspection, the translation process generally keeps the cellular arrangement and structure of the source image while keeping the contents compatible with the target tumor type. For example, colorectal adenocarcinoma (COAD) usually features the glandular structures formed by the epithelial cells (**Fig. 3b**). When the colorectal images are translated to other adenocarcinomas, those structures are kept intact (**Fig. 3b**). However, when they are translated to non-adenocarcinomas, those structures are attenuated as much as possible and replaced with some target type-specific contents (**Fig. 3b**).

**Figure 3.**
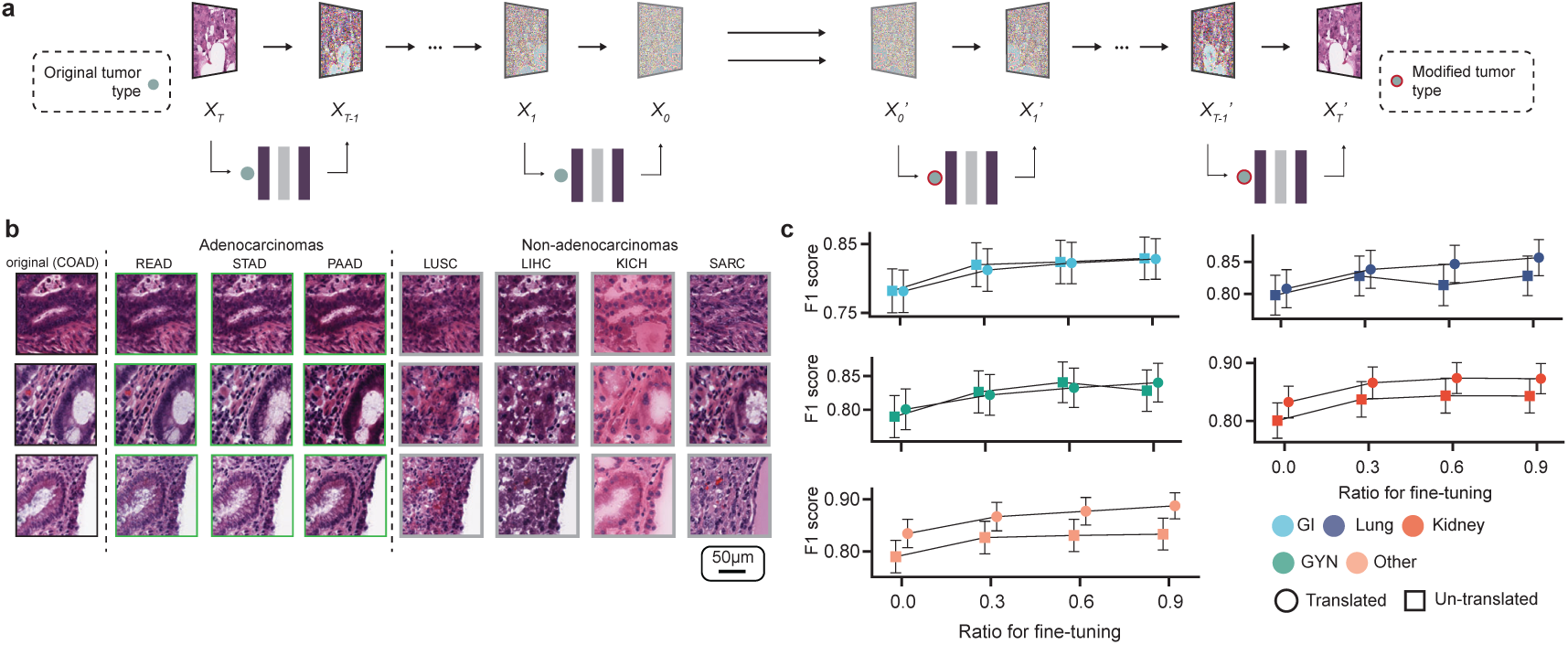
Translation of histopathological images across primary tumor types. (a) Schematic that illustrates how His-MMDM translates across primary tumor types. (b) Image examples showing the effect of translation across tumor types. The COAD patches were translated across three adenocarcinomas (READ, STAD, and PAAD) and four non-adenocarcinomas (LUSC, LIHC, KICH, and SARC). The typical glandular structures of adenocarcinomas were retained in the former group but attenuated in the latter group. (c) The classification performance (F1 score) of binary tumor classifiers that are trained on other tumor types (as targets of translation) using either translated/untranslated images when there is no additional fine-tuning (‘Ratio for fine-tuning = 0’) or with additional fine-tuning (‘ratio for fine-tuning = 0.3, 0.6, 0.9’) in each tumor type (as the source of translation and the goal of classification). Data for each source of translation are aggregated for a particular tumor family and through macro-averaging.

Translation of images from one tumor type to another enables new applications. For example, suppose we have a discriminative model for tumor type A and we would like to apply it to another tumor type B where such a model is unavailable. Instead of directly applying it to the images from B, which is the cross-classification strategy as in Noorbakhsh et al. ^42^, we could first adapt the images into type A and then use the model to classify or further fine-tune the model on the adapted images. In this way, we could use His-MMDM to achieve *knowledge transfer* across tumor types. We compared the performance (in terms of F1 score) of binary tumor classification models trained on one tumor type (the target of translation) and cross-classifying images from another tumor type (the source of translation/the goal of classification) using either the original images or the translated images. The classification performance in four out of five tumor families corresponding to 12 out of 19 tumor types on average showed higher performance using the translated images (**Fig. 3c** and **Fig. S2c-S2d**, ‘ratio for fine-tuning = 0’). Additionally, to improve the classification models’ performance, we fine-tuned the classification models using 30%, 60%, and 90% of the translated images (**Fig. 3c** and **Fig. S2c-S2d**, ‘ratio for fine-tuning = 0.3, 0.6, 0.9’). The number of tumor types with higher performance using the translated images increases to 13 at 60% and 15 at 90%.

### Knowledge transfer of histopathological images between primary and metastatic organ sites

Metastasis of tumor is one of the leading factors that reduces a patient’s life quality and survival. Therefore, the prediction of metastasis of tumors has important application values in clinical diagnosis. From a histopathological perspective, tumors displaying higher levels of malignancy tend to have a higher tendency to metastasize ^44^. Several previous studies have already attempted to build discriminative models that can infer the metastatic potential of tumors from such images ^45–47^.

Using His-MMDM, we investigated our model’s potential application on this task but from a generative model’s perspective. Again, we aim to address the following question about *knowledge transfer*: can His-MMDM leverage the knowledge it has acquired from generating images of metastatic organ sites to provide prognostic and staging capabilities for primary site images? We trained His-MMDM on 475 lung tumor histopathological WSIs, including 315 from primary lung tumors, 76 from lymph node metastases, and 84 from brain metastases from the Harbin Medical University Cancer Hospital (the ‘HMU-C dataset’, **Methods**, **Table S2**). During training, His-MMDM was conditioned on the site (lung, lymph, and brain) of the tumor tissue, and during inference, His-MMDM was asked to translate the images from one site to another (**Fig. S3a**). As before, we computed ΔFID between each pair of translations. The ΔFID scores are much higher when translating from a non-solid organ site (lymph node) to a solid organ site (lung and brain) and the translation within solid organ sites (lung and brain) tends to result in lower ΔFIDs (**Fig. 4a**). Inspecting the original and translated histopathological images, His-MMDM keeps the cellular arrangements and characteristics of the tumor subtypes during the translation processes, but the tumor growth environment is modified to match the designated organ sites (**Fig. 4b**).

**Figure 4.**
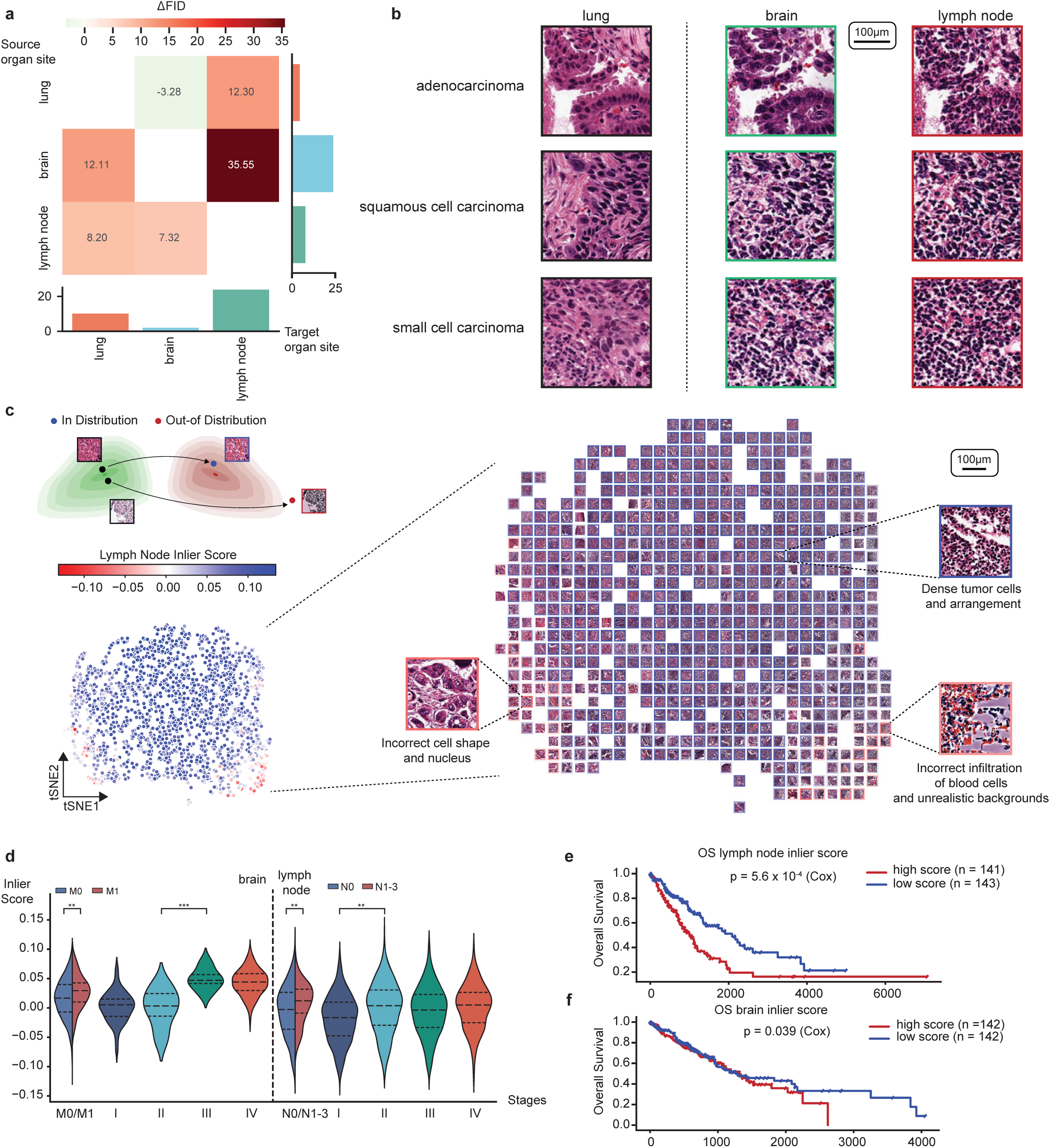
Translation of histopathological images between primary and metastatic organ sites. (a) The improvement of FID score before and after translation between each pair of organ sites (lung, lymph node, and brain). Bar plots show the averaged ΔFID for a particular translation source site (rows) or target site (columns). (b) Histopathological image examples of three major lung tumor subtypes (adenocarcinoma, squamous cell carcinoma, and small cell carcinoma) illustrating the effect of translating images from the primary site (lung) to the common metastatic sites of lung tumor (brain and lymph node). (c) Schematic and visualization of running the outlier detection model (Isolation Forest) on the translated images from the lung to the lymph nodes. The images are visualized according to their structure in the tSNE space and the ‘inlier scores’ are superimposed. The images with low ‘inlier scores’ (out of distribution) tend to be eccentric-looking and are located on the edge. (d) The relationship between the brain inlier score and the stagings of cases in the TCGA lung tumor cohort. The stagings include the M (distant metastasis) and N (extent of regional lymph node spread) stagings in the TNM staging system. (e-f) The relationship between the lymph node (e) and brain (f) inlier score to TCGA patients’ overall survival. The inlier scores of the images are aggregated per patient through averaging.

We systematically visualized the images translated from the primary organ site (lung) to the metastatic organ sites, including the lymph node (**Fig. 4c**) and brain (**Fig. S3b**) according to their Inception V3 features in the 2D tSNE space. The images located at the interior of the distribution are more likely to be properly translated from the primary site characterized by the properly arranged tumor cell nuclei and the more realistic background tumor environment when assessed by pathologists (**Fig. 4c**, **Fig. S3b** and **Fig. S3c**). On the contrary, the translated images that look eccentric (characterized by incorrect cell shape, infiltration, and unrealistic backgrounds) tend to be located at the edge of the distribution whose appearance indicates less successful image translation which could be attributed to a lack of compatibility of the primary site image with the metastatic site (**Fig. 4c**). We trained an outlier detection model, Isolation Forest ^48^, on the Inception V3 features of the translated images and produced an ‘inlier score’ (in the range [-1,1], –1 for the most out-of-distribution and 1 for the most in-distribution) for each lung image to measure its compatibility with the metastatic organ site (**Fig. 4c** and **Fig. S3b**). We speculated tumors that are easy to translate to a particular metastatic site image will have a higher ‘inlier score’ (compatibility) with that particular organ site. To verify this, we translated primary lung tumor histopathological images (LUAD and LUSC) from TCGA to the metastatic organ sites (brain and lymph node) using the trained His-MMDM model and computed their inlier scores using the outlier detection model. Interestingly, we found that the inlier scores indeed displayed associations with the staging of the patients. Specifically, a patient’s high brain inlier score is associated with M1 staging and an overall staging of III-IV, and a patient’s high lymph node inlier score is associated with N1-N3 staging and an overall staging of II-IV (**Fig. 4d**). Moreover, a high lymph node inlier score is associated with poorer survival of the patients (**Fig. 4e**, p=5.6e-04). This trend also holds for the brain inlier score (**Fig. 4f**, p=0.039).

### Genomics– and transcriptomics-guided editing of histopathological images

Somatic mutations in oncogenes and tumor suppressors are the driver factors in tumorigenesis. Previous studies have reported strong associations of driver mutations with histopathological image features, which were demonstrated through the predictive power of discriminative models ^49–55^. More recently, pan-cancer studies have shown the identifiability of common driver mutations (e.g. *TP53* and *PTEN*) shared in multiple tumor types using deep transfer learning ^56^. Apart from genomic mutations, several other works have been dedicated to the prediction of transcriptomic expression levels from histopathological images, achieving various levels of success ^56,57^.

We explored the capability of His-MMDM to decipher these associations but from a generative model’s perspective. To this end, we trained another His-MMDM model from the TCGA histopathological images conditioned simultaneously on the genomic mutation profiles and the transcriptomic profiles of the corresponding samples (**Methods**). In total, the genomic embeddings of His-MMDM take into account the 334 genes from the ten common oncological signaling pathways ^58^, and 188 other genes with high pan-cancer mutation rate or discriminative performance ^56^ in the TCGA cohort (**Table S5**). Similarly, the transcriptomic embeddings include 4,193 genes that were included in the collection of 50 hallmark pathways of MSigDB (v2023.2) ^59,60^, as well as an additional 268 genes that either displayed the greatest variability between tumor and normal samples or have previously reported discriminative performance ^57^ (**Table S5**).

We first performed *in silico* genomic modification experiments using the trained His-MMDM model. For a given histopathological image, the forward diffusion process is executed upon it conditioned on its original genomic mutation profile, e.g., gene X is wide-type (WT) and gene Y is mutated, and the backward denoising process is then executed conditioned on a modified genomic profile, e.g., gene X is mutated and gene Y is WT (**Fig. 5a**). In this way, we are essentially asking the model to edit the given histopathological image into a version if its gene X and gene Y’s mutation statuses were as designated.

**Figure 5.**
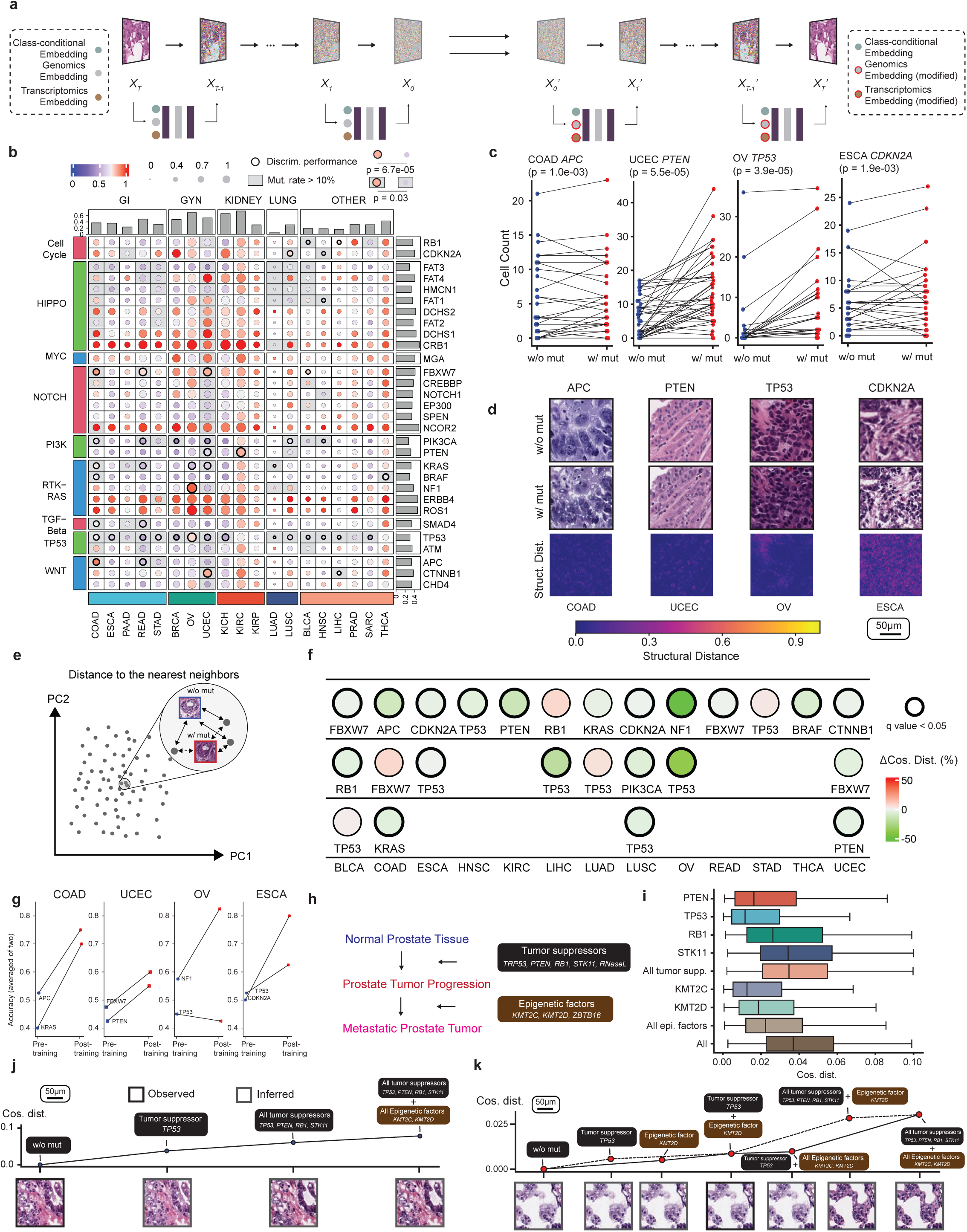
The genomics-guided editing of histopathological images. (a) Schematic that illustrates how His-MMDM achieves histopathological image editing guided by genomic and transcriptomic profiles. (b) The effect of genomic mutations on histopathological images by each tumor type, measured by cosine distance between the WT version image and mutated image. The top 30 genes with the highest pan-cancer mutation rate are selected. The cosine distances aggregated by each gene-tumor type pair through averaging are normalized either by each tumor type (circle color) or cross-tumor (circle size). Bar plots aggregate per each gene (row) or tumor type (row). The mutations of genes have a greater effect on histopathology in the gene-tumor type pairs where there were previously reported associations by Fu et al. (Wilcoxon rank-sum test; all gene-tumor pairs: p = 6.7e-05; only high-frequency (>10%) mutations: p = 0.03). (c) The mutation of genes *APC*, *PTEN*, *TP53*, and *CDKN2A* increases the numbers of malignant cells detected by Hover-Net in COAD, UCEC, OV, and ESCA, respectively. P values are from the Wilcoxon signed-rank test. Each dot represents a particular image without (blue) or with (red) a particular mutation. (d) Histopathological image examples illustrating the effect of mutations of *APC*, *PTEN*, *TP53*, and *CDKN2A* in COAD, UCEC, OV, and ESCA, respectively. Structural distance maps are shown in the last row to highlight the differences between the two versions. (e-f) The computation of the change in cosine distance of a query image (w/o mut) to its nearest neighbors in the database when the image is edited into a version with mutation (w/mut) is illustrated in (e). 18 out of the top 25 tumor type and mutation pairs where His-MMDM’s edits resulted in the most notable feature changes displayed a statistically significant reduction in the distance to their nearest neighbors (f). q values are from the Wilcoxon signed-rank test adjusted by the Benjamini–Hochberg procedure. (g) Performance of pathologists’ recognition of the top genomic mutations in four tumors (COAD, UCEC, OV, and ESCA) before and after observing His-MMDM’s generated images as a tutorial. (h) Summary of the effect of five tumor suppressors and three epigenetic factors on the development and metastasis of prostate tumors reported by Cai et al. (i) The effect of genomic mutations in the tumor suppressors and epigenetic factors on the histopathology images. (j) An example illustrating His-MMDM’s ability to infer images at different tumor developmental stages (mutation in one tumor suppressor gene, mutation in all tumor suppressor genes, and mutation in all tumor suppressor and epigenetic factor genes) from a normal image. Cosine distances are computed w.r.t. the initial normal image (w/o mutation). (k) An example illustrating His-MMDM’s ability to infer images when there is more mutation or less mutation than the observed image (mutation in *TP53* and *KMT2D*). Such inference can be made along different paths, i.e., the solid line and the dashed line, representing mutations accumulating in different orders. Cosine distances are computed w.r.t. the initial inferred image (w/o mutation).

We first investigated the effect of single mutations among the top 30 genes with the highest mutation rates across the aforementioned 19 tumor types on the appearance of histopathological images. For each image, we used His-MMDM to produce one version of it when all genes are WT. Then we produced a series of other images when each one of the genes is mutated. The mutation of different genes resulted in modifications in different aspects of the histopathological image. For example, in COAD, the mutation of *APC* had the most significant effect on the nuclear shape of the cell, *TP53* on the cellular density, and *SMAD4* on the background and microenvironment (**Fig. S4a**). To measure the visual effect of the mutation, we computed the cosine distance between the WT image and each ‘mutated image’. Specifically, for each gene in a specific tumor type, the cosine distances of the Inception V3 features of the two versions are averaged and normalized both within– and cross-tumor (**Fig. 5b**). Some tumor types show the greatest overall effect of single mutations on their visual appearance, such as the kidney tumors (KICH, KIRC, and KIRP). While some other tumor types showed greater effect within their tumor categories, such as COAD and READ among the GI tumors and OV in GYN tumors (**Fig. 5b**). Mutations in HIPPO, NOTCH, and RTK-RAS pathways tend to have the greatest pan-cancer effect on histopathology. Interestingly, these genes in NOTCH and RTK-RAS are *NCOR2* and *ERBB4*, but are not *NOTCH1* and *KRAS* themselves (**Fig. 5b**). The mutations of genes have a greater effect on histopathology in tumor types where there were previously reported associations between them based on discriminative performance reported by Fu et al. ^56^ (all gene-tumor pairs: p = 6.7e-05; p = 0.03 only high-frequency mutations (>10%)). We repeated the experiments using structural distance (**Methods**) instead of InceptionV3 cosine distance and a very high correlation was observed between the two (**Fig. S4b-S4c**). We applied the cell detection algorithm, Hover-Net ^61^, to the generated images of the genes which have high mutation rates across multiple tumors. Most notably, the generated images corresponding to the mutated version of *APC*, *PTEN*, *TP53*, and *CDKN2A* have a higher number of malignant cells compared to their WT counterparts in COAD, UCEC, OV, and ESCA, respectively (**Fig. 5c and 5d**). Hereafter, a structural distance map for each edited image to highlight the edited regions (**Fig. 5d**, **Methods**).

We further validated the results of genomics-guided image editing by searching both the original and edited images in a curated histopathological image database with accompanying genomics profiling data (**Methods**). The database is curated to include real histopathological images that represent various genomic mutations across different tumor types. We compared the original images (w/o mutation) and edited images (w/ mutation), calculating the cosine distance (InceptionV3 feature) between each image and its nearest neighbor in the database (**Fig. 5e**). For the top 25 tumor type and mutation pairs where His-MMDM’s edits resulted in the most notable feature changes (**Fig. 5b**), 18 pairs showed a statistically significant reduction in the distance to their nearest neighbors (**Fig. 5f**). Additionally, we used these paired images (w/ and w/o mutation) as educational material for pathologists, assessing their ability to recognize such mutations after reviewing these examples (**Methods**, **Supplementary Data**). We observed an improvement in the pathologists’ accuracy following exposure to these examples of seven out of eight mutations in the four tumor types COAD, UCEC, OV, and ESCA (**Fig. 5g, Fig. S4d-S4e**). These findings suggest that the image content generated by His-MMDM aligns well with reality.

Apart from single mutations, we also investigated the effect of accumulating mutations on the generated histopathological images by His-MMDM. For a total of 331 genes in nine oncological pathways (one pathway excluded due to its low number of genes), we sorted the genes in each of them according to their mutation rate (from high to low, **Table S4**) and edited the images by sequentially performing the mutation of 25%, 50%, 75%, and 100% genes in each pathway. We then compared the cosine similarity of Inception V3 features of each of them to the WT version of the image (**Fig. S4f**). Some pathways, such as Cell Cycle and WNT signaling pathways in COAD, displayed continuous changes as the mutation accumulates (**Fig. S4f, S4g**). But more frequently, a saturation effect was observed after the most significant change had already resulted from the initial 25% of the mutations (**Fig. S4f**).

Using a CRISPR/Cas9 mouse model, a recent study ^62^ has confirmed the sufficiency of loss-of-function mutations of five tumor suppressor genes (*PTEN*, *TRP53* (mouse homolog of the human *TP53*), *RB1*, *STK11*, and *RNaseL*) to induce prostate tumor progression and three additional epigenetic factors (*KMT2C*, *KMT2D*, and *ZBTB16*) to initiate tumor metastasis (**Fig. 5h**). Among them, only *RNaseL* and *ZBTB16* are not included in the His-MMDM genomic embeddings. We therefore attempted to edit the PRAD images in the TCGA cohort according to the mutation of the remaining seven genes. For each image, we used His-MMDM to produce one version free of mutations in all tumor suppressors and epigenetic factors (WT), and several mutant versions for (1) individual mutations in the tumor suppressors and epigenetic factors, (2) combined mutations in all tumor suppressors, (3) combined mutations in all epigenetic factors, and (4) combined mutations in all tumor suppressors + epigenetic factors. We then computed the cosine distances of the Inception V3 features of the mutated images versus the WT images (**Fig. 5i**). Among the tumor suppressors, *STK11* induces the greatest histopathological changes in the tissue histopathology (**Fig. 5i**). The tumor suppressors generally had higher impact than the epigenetic factors, which is consistent the previous study that it is these factors that mainly contributed to the *in situ* tumorigenesis of prostate tumor. The advantages of His-MMDM enabled *in silico* interpolation of histopathological images corresponding to different mutation statuses of the tumor. As for images from normal tissue, His-MMDM can infer its appearance when one or more of the tumor suppressors/epigenetic factors are mutated (**Fig. 5j**). As for images from tumor tissue with some somatic mutations, His-MMDM can traceback to its WT appearance or infer later stages of the tumor with more accumulating mutations. For instance, for an image from the prostate tumor with existing mutations in *TP53* and *KMT2D*, His-MMDM can edit it into more or fewer mutations in the tumor suppressors/epigenetic factors and can freely modify it into versions when the mutations of tumor suppressors and epigenetic factors are applied either together or alone (**Fig. 5k**).

Besides genomic mutations, we investigated the effect of modifying transcriptomic profiles on the generated histopathological images. As transcriptomic profiles are shaped by regulations of pathways, we first investigated the effect of the top 10 MSigDB transcriptomic pathways that are most dysregulated in TCGA tumor samples (**Methods**). To exaggerate the effect of transcriptomics on the generated images, for each histopathological image in the TCGA cohort, we used His-MMDM to produce one version of it corresponding to when a pathway was knocked up and another one when it was knocked down. Similarly, we computed the cosine distance (**Fig. 6a**) and structural distance (**Fig. S5a**) between the two versions of the image and visualized it accordingly. Most notably, the manipulation of the ‘Mitotic Spindle’ pathway resulted in the greatest change in histopathology in almost all tumor types (**Fig. 6a** and **Fig. S5a**). The result is not surprising as tumor proliferation through mitosis usually has a high correlation with histopathological grading. Targets of the transcription factor Myc (‘Myc Targets V1’) are influential in the tumorigenesis of multiple GI tumors, and therefore, they display a high impact on the histopathology of COAD, READ, and STAD (**Fig. 6a** and **Fig. S5a-S5b**). The manipulation of transcriptomic pathways also had a greater histopathological effect in tumor types when their genes had previously reported associations with histopathology based on discriminative performance reported by Fu et al. ^56^ (p=5.2e-03). Similar to genomics-guided editing, 20 out of the top 25 tumor type and pathway pairs displayed a statistically significant reduction in the distance to their nearest neighbors in the database curated for transcriptomics pathway up-regulation (**Fig. S5c**, **Methods**). Using these images as educational materials, we also observed an improvement in the pathologists’ accuracy following exposure to these examples (**Fig. S5d**). Using Hover-Net, we observed a significant reduction in the number of malignant cells when either the ‘G2M checkpoint’ (in 9/19 tumor types) or the ‘DNA repair’ (in 7/19 tumor types) pathways was knocked up compared to when it was knocked down (**Fig. 6b-c, Fig. S5e** and **S5f**), suggesting that His-MMDM can indeed capture the effect of the activation of such pathways in limiting tumor growth potential.

**Figure 6.**
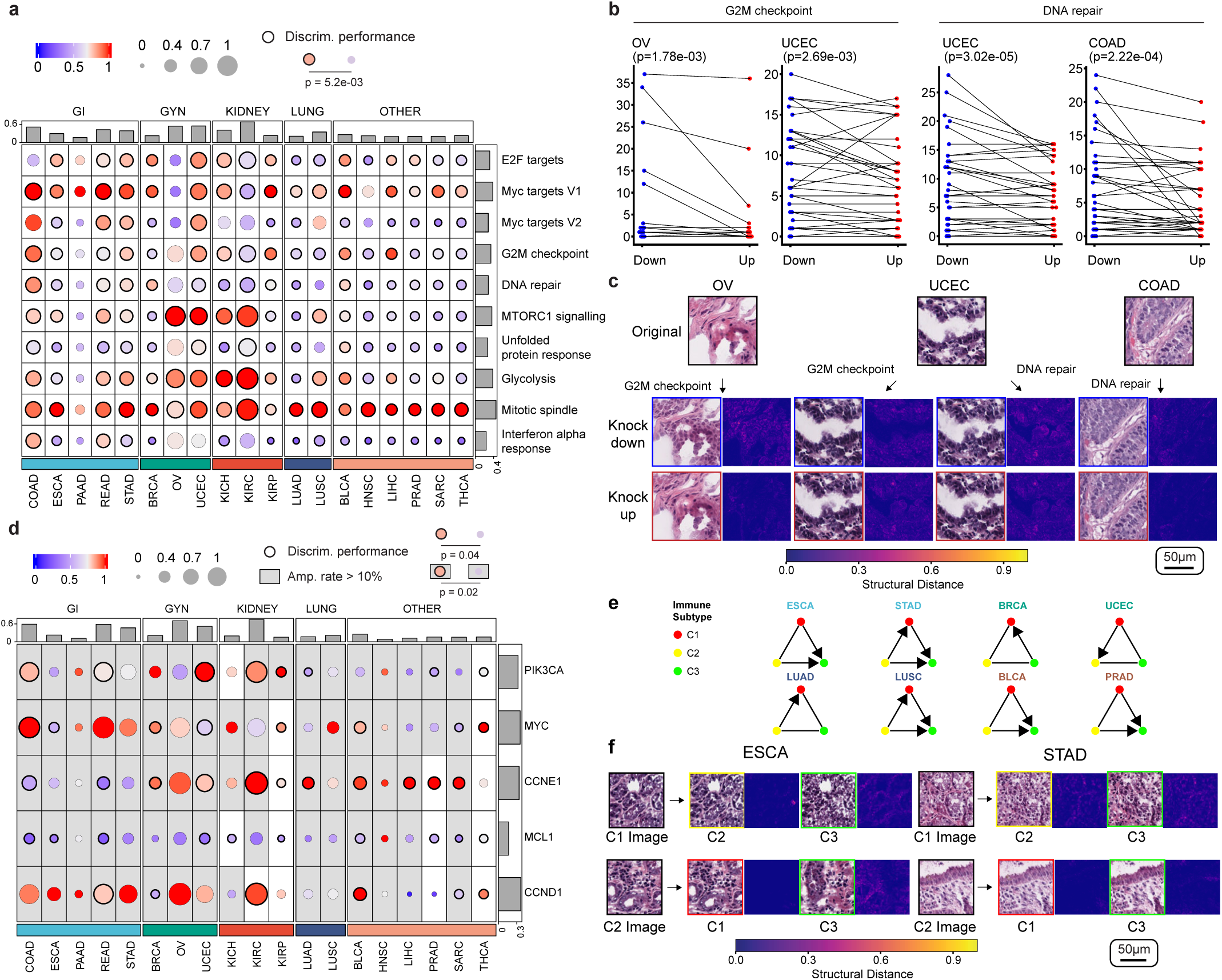
The Transcriptomics-guided editing of histopathological images. (a) The effect of transcriptomic pathway manipulations on the histopathological images by each tumor type, measured by the cosine distances between a knocked-down version and a knocked-up version. The top 10 pathways with the highest t-statistics between TCGA tumor and normal samples are selected for the experiment. The cosine distances aggregated by each pathway-tumor type pair through averaging are normalized either by each tumor type (circle color) or cross-tumor (circle size). Bar plots aggregate per each pathway (row) or tumor type (row). The manipulation of pathways would have a greater effect on histopathology if the pathways have genes with previously reported histopathological associations by Fu et al. (Wilcoxon rank-sum test; p = 5.2e-03). (b) The knock-up of the pathway ‘G2M checkpoint’ reduces the number of malignant cells detected by Hover-Net in OV and UCEC respectively. The knock-up of the pathway ‘DNA repair’ reduces the number of malignant cells detected by Hover-Net in UCEC and COAD respectively. Each dot represents a particular image when a particular pathway is knocked down (blue) or knocked up (red). (c) Histopathological image examples illustrating the effect of knock-up and knock-down of the ‘G2M checkpoint’ and the ‘DNA repair’ pathways in OV, UCEC, and COAD. Structural distance maps are shown to highlight the differences. (d) The effect of transcriptomic manipulations of high-frequency SCNV genes reported by Aaltonen et al., measured by the cosine distances between a knocked-down version and a knocked-up version. Bar plots aggregate per each gene (row) or tumor type (row). The manipulation of genes would have a greater effect on histopathology if the genes have previously reported histopathological associations by Fu et al. (Wilcoxon rank-sum test; all gene-tumor pairs: p = 0.04; only genes with high amplification rates (>10%): p = 0.02). (e) Editing of histopathological images based on transcriptomic signatures of immune profiles that result in a significantly higher number of necrotic cells detected by Hover-Net. An arrow is drawn from one immune subtype to another if the increase of such cells is statistically significant (Wilcoxon signed-rank test, p < 0.001). (f) Histopathological image examples illustrating the effect of editing images into different immune subtypes from their original ones.

Aside from pathway-level alterations, transcriptional changes in individual genes could also be critical in tumorigenesis. Most notably, the amplification of genomic segments can directly alter the transcriptomic expression level of genes through somatic copy number variations (SCNVs). We investigated the five genes with high pan-cancer SCNV rates reported by Aaltonen et al. ^63^ that are also available in the transcriptomic embeddings of His-MMDM. These include the prominent oncogenic transcription factor, *MYC*, a core component of the PI3K signaling pathway, *PIK3CA*, two genes from the cyclin family, *CCNE1* and *CCND1*, and one gene from the *Bcl-2* family, *MCL1*. We used His-MMDM to produce one image with an average expression level of the genes in the normal samples and compared it against another version when each of their expression levels was elevated (3 × s.d. above the average). Most notably, we observed a greater effect of the cyclin genes, *CCNE1* and *CCND1,* on the tumor histopathology, suggesting the SCNVs that promote the progression of the cell cycle may have a universal effect across tumor types (**Fig. 6d, Fig. S5g**). In line with the results of Myc target genes in the pathway analysis, the gene *MYC* itself also induced strong histopathological alterations in multiple GI tumor types (**Fig. 6d, Fig. S5g**). As before, for these SCNV genes, there is also an association between their greater histopathological effect and previously reported discriminative performance on their transcriptomic levels (p=0.04 overall and p=0.02 if only tumor types where the genes have high amplification rates are considered).

The immune microenvironment of tumors plays an essential role in tumor development, subtyping, and prognosis ^64^. Thorsson et al. systematically characterized the immunogenomic signatures in TCGA and classified each tumor sample into six immune subtypes (C1-C6) based on the clustering of transcriptomic profiles of key immune pathways ^64^. As it is reported that tumor samples in such immune subtypes display distinct characteristics such as tumor proliferation and the types of infiltrated immune cells, we were interested in investigating whether His-MMDM can reproduce such observations. We selected the most abundant three (C1, C2, and C3) out of the six immune subtypes studied by Thorsson et al. and dropped the three kidney tumor types due to their low numbers of C1 and C2 immune subtypes (see Fig. 1D of Thorsson et al. ^64^). We then used His-MMDM to edit the images from one immune subtype into another one (e.g. editing C1 images into C2 or C3) by manipulating the five underlying pathways that define the immune subtypes (**Methods**, **Table S5**). The detection of cell types using Hover-Net on the images before and after His-MMDM’s edit revealed an increase (with a few exceptions) in the number of necrotic cells when translating images from C1 to C3 (in five tumor types) and C2 to C3 (in five tumor types) (**Fig. 6e**-**f**, **Fig. S5h**) which is consistent with C3’s lower tumor cell proliferation and high inflammatory response. As a negative control, we used randomly selected sets of genes (while keeping the sizes of the gene sets the same) (**Table S5**) for the underlying immune pathways and did not observe statistically significant increments in most cases (**Fig. S5i**). We finally demonstrate through an example when His-MMDM is guided by the combination of genomic mutations and transcriptomic profiles to edit histopathological images between BRAF-like and RAS-like thyroid tumors. However, we delegate such discussions to the Supplementary Materials (**Section S3**, **Fig. S5j-S5k**).

## Discussion

In this study, we have developed His-MMDM, a histopathological image translation model that translates histopathological images between multiple categorial domains or edits them guided by genomic or transcriptomic profiles. A lot of previous evidence has suggested the feasibility of building such general image translation models across histopathological domains. Firstly, several previous studies have pointed out the existence of conserved histopathological image features across different tumor types ^42,43^. His-MMDM also demonstrates this via explicitly translating images and transferring knowledge between them. Secondly, pan-cancer studies have demonstrated through discriminative models the high predictive performance of the tumor origin from histopathological images ^65^, multiple genomic mutations ^56^, and multiple genes’ expression levels ^57^. We demonstrated, for the first time, that such relationships can be manifested through a generative model, by explicitly generating histopathological images corresponding to *in silico* altered tumor types or genomic and transcriptomic profiles. Thirdly, several dedicated histopathological image translation models have already been proposed in specific application domains such as cryosection to FFPE translation and *in silico* virtual staining, which proves the applicability of image translation in histopathology ^25,26^. We have shown that the general His-MMDM model can achieve performance comparable to these dedicated models. Most notably, we illustrated that such translation from cryosection to FFPE not only improves the visual quality for pathologists, but is useful to improve the performance of existing histopathological foundation models as well.

Very recently, several new generative models to synthesize histopathological images based on genomics or transcriptomics have begun to emerge. For instance, Dolezal et al. proposed a conditional GAN (cGAN) based model ^66^ that can synthesize images of different subtypes in lung, breast, head & neck, and thyroid tumors. In particular, the model can synthesize thyroid tumor images from BRAF^V600E^-like ones to RAS-like ones. Carrillo-Perez et al. sequentially developed a GAN-based ^67^ and a diffusion-based generative model ^68^ for the synthesis of histopathological images from bulk RNA-seq profiles. It is worth noting that these previous works are generative models conditioned only on a genomic/transcriptomic profile and synthesize new images *from scratch*. His-MMDM, however, distinguishes itself as an image translation model that can edit existing images into new ones. This property is indispensable in our analyses of the effect of genomic/transcriptomic manipulations on real histopathological images. This also makes His-MMDM more useful in real-world applications in that it achieves multi-omics-guided editing of the existing images rather than the un-constrained generation of new ones.

In recent years, GenAI-based copilot systems have shown great potential to revolutionize the way we interact with computers ^69–71^. In parallel, preliminary attempts have also been taking place in biomedicine and such GenAI systems are showing great potential in helping biologists and healthcare practitioners ^19,20,72,73^. As a GenAI model itself, His-MMDM could fit naturally into such ecosystems and become useful for a pathologist’s day-to-day workflow. One limitation of His-MMDM concerns its high computational demand. Due to the iterative nature of the diffusing and denoising processes of diffusion models, it still takes minutes to translate a single batch of histopathological image patches on a typical machine equipped with eight NVIDIA V100 GPUs. Strategies for developing more efficient diffusion models ^74^ can be adopted to streamline the process of image translation. Experiments with more efficient diffusion model solvers ^75,76^ can also be an interesting exploration for future works. Currently, for simplicity, we trained His-MMDM with fixed resolution (128×128) under a unified magnification level in each cohort. Extending His-MMDM to simultaneously work at different magnification levels will further unleash its application potential.

## Methods

### Datasets

In this research, three cohorts of histopathological image datasets were used to train and test the His-MMDM model under different image translation settings.

*The Cancer Genome Atlas cohort (TCGA)* We downloaded 22,596 whole-side histopathological images from The Cancer Genome Atlas (TCGA) web portal (https://portal.gdc.cancer.gov/) of the 19 cancer types following the practice of ^42^. The 19 cancer types contain five gastrointestinal (ESCA, STAD, COAD, READ, and PAAD), two lung (LUAD and LUSC), three kidney (KIRP, KICH, and KIRC), three pan-gynecological (OV, BRCA, and UCEC), and five other tumor types (LIHC, HNSC, SARC, THCA, BLCA, and PRAD) (statistics shown in **Table S2**). These tumor types in TCGA were specially chosen because they contain enough numbers of tumor and normal slides, as well as enough numbers of FFPE and cryosectioned slides for analyses. The patient’s metadata and clinical information were downloaded along with the WSIs. The processed genomic mutations (TCGA Unified Ensemble “MC3” gene-level mutation calls) and the bulk transcriptomic profiles (Illumina Hi-Seq) of the corresponding samples were downloaded from the UCSC Xena website (https://tcga.xenahubs.net) ^77^. This cohort is used for training and testing His-MMDM to translate across different primary tumor types and different genomic and transcriptomic profiles. For genomic profiles, the 522 genes we considered in total include the 334 genes from the ten oncological signaling pathways defined in ^58^, and 188 other genes with high pan-cancer mutation rate or discriminative performance ^56^ in the TCGA cohort. We strictly followed the “MC3” gene-level mutation calls’ definition and only non-synonymous mutation statuses were considered for each gene. For transcriptomic profiles, we considered a total of 4,461 genes that are available in the TCGA transcriptomic profiles, including 4,193 genes that are included in the collection of 50 hallmark pathways of MSigDB (v2023.2) ^59,60^, as well as an additional 268 genes that either displayed the greatest variability between tumor and normal samples (in terms of the t-statistic) or previously reported discriminative performance ^56^. During the evaluation of the His-MMDM model, we prioritized the top 30 genes in terms of pan-cancer mutation rate from nine oncological pathways for genomic-guided editing and the top 10 pathways in terms of pan-cancer t-statistic from MSigDB for transcriptomic manipulation.

*The Harbin Medical University Cancer Hospital Cohort (HMU-C)* This cohort contains 475 primary and metastatic lung tumor histopathological WSIs of 400 patients from the Harbin Medical University Cancer Hospital. This HMU-C cohort consists of 315 WSIs from primary lung tumor tissues, 76 WSIs from lymph node metastases, and 84 WSIs from the brain metastases of the same set of patients. The primary and metastatic tumor tissues are from the most common lung tumor subtypes: adenocarcinoma, squamous cell carcinoma, and small cell carcinoma (statistics in **Table S2**). All slides in this cohort are FFPE slides.

*The First Affiliated Hospital of Harbin Medical University Cohort (HMU-1st)* This cohort contains 6,200 brain tumor histopathological WSIs of 557 patients from the First Harbin Medical University Hospital. Among the 6,200 WSIs, 2,660 are glioma slides, and 3,540 are meningioma slides; 2,753 were H&E-stained and 3,447 were IHC-stained with 14 different markers (**Table S4**). Some of these markers are useful for the confirmation of the source of the tumor tissue, such as ATRX; some are used to display specific cellular processes, such as Ki67; while others are useful in tumor subtyping and forecasting prognoses, such as IDH-1 and MGMT (**Table S4**). Among them six markers are commonly used in both meningiomas and gliomas, four are used exclusively in gliomas and the other four are used exclusively in meningiomas (**Table S4**).

The protocols for the human studies comply with all relevant ethical regulations and are approved by the Ethics Committee of The First Harbin Medical University Hospital and The Harbin Medical University Cancer Hospital (KY2021-42). The consent forms of the patients were waived before this research was carried out under the retrospective research protocol of the institutions.

### Patch selection and patch feature extraction

The WSIs from each cohort were segmented for tissue regions from the empty slide background. The slide image was then converted to a binary mask using Otsu’s thresholding method on the Gaussian blurred version of the saturation channel. Subsequently, using a sliding window-based approach, the carved-out tissue area is completely covered with image patches that are 256 × 256 (if the objective magnification is 20 ×) or 512 × 512 (if the objective magnification is 40 ×) in size. To select patches from the WSIs that contain representative tumor tissues, we extracted the image patches’ features using a pretrained (on ImageNet ^78^) ViT-L-16 ^79^ model. From each cohort’s extracted patches, we asked the pathologists to select a small set of positive patches (which contains representative tumor cells and microenvironment) and a small set of negative patches (in which the tumor tissue content is less than 10% of the area covered). We then classified the patches as positive/negative from each WSI based on their similarity to the positive/negative patch sets (as measured by the cosine similarity of their ViT-L-16 feature vectors). From each WSI, we randomly sampled a certain number of patches from the ‘positive patches’ determined above and then incorporated them in the train/test set of the His-MMDM model. The patches were resized to 128×128 before being processed by the generative models. The statistics of the extracted patches in each cohort are shown in **Table S2**.

### The Multi-domain multi-omics image translation model

His-MMDM is an image translation model that learns a mapping 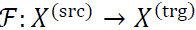 that maps a source domain image 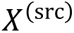 to a target domain image 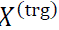. If the images in the source domain are distributed as 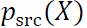 and the images in the target domain are distributed as 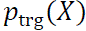, a well-trained image translation model is expected to have the property that 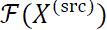 distributes as 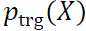. Although it is possible to translate images between different dimensions ^30^, we are assuming that 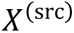 and 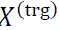 to have the same dimensions in this paper for simplicity. This assumption is valid since we have resized images to represent the same physical dimension when they come from slides with different magnification levels.

Su et al. have demonstrated theoretically that such mapping ℱ corresponds to a deterministic solution of the probability flow ordinary differential equations (PF-ODEs) of the Schrödinger Bridge Problem (SBP) between the two distributions *p*_src_ and *p*_trg_ ^30^. The SBP from *p*_src_ to *p*_trg_ aims to establish the most likely evolutionary path of a distribution from *p*_src_ to *p*_trg_. They also established the connection between the SBP and the score-based generative modeling (SGM) of DMs and proved that SGM is a special case of the SBP when the evolution is from a particular data distribution (could be *p*_src_ or *p*_trg_) to the multivariate Gaussian distribution. In this way, such mapping ℱ can be found by composing the forward (from 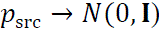) and backward diffusion processes (from 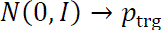) of the SGM. Concretely, during the forward diffusion processes, we obtain the latent noisy image 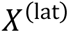 from a source domain image 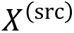 with

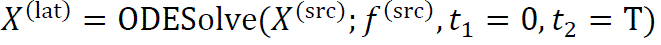

where 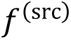 is the denoising diffusion model trained in the SGM of the source domain, ODESolve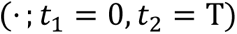 represents the numerical approximation of the *forward* PF-ODE that maps the source domain image 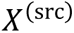 to the latent image 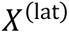, and T is the preset number of discretization steps of the solver. If we assume that the SGM models perfectly the source domain score function and there is no discretization error in solving the PF-ODE, 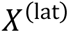 should distribute as *N*(0, **I**). Symmetrically, we can then transform the latent image 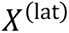 to the target domain with

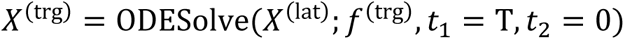

where 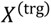 is the generated target domain image, ODESolve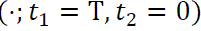 represents the numerical approximation of the *backward* PF-ODE that maps the latent image to the target domain image, and 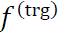 is the denoising diffusion model of the target domain SGM.

His-MMDM focuses on the translation of histopathological images belonging to different domains. Such domains include categorical domains such as different organ sites of the tumor and different types of stains, as well as multi-omic domains that correspond to different genomic or transcriptomic profiles. Therefore, the denoising diffusion model used in His-MMDM needs to simultaneously take care of the three types of condition information. For clarity, we first write the categorical condition as *c* ∈ *C*, where *C* is a finite set containing all the possible categorical conditions that are handled by this model. Secondly, we use 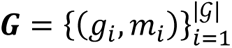 to hold the genomic mutation profiles of all the genes considered by the model (denoted by gene set *G*) for a particular train/test example, in which *g*_*i*_ is the name of the *i*th gene and *m*_*i*_ is the Boolean value of its mutation status. Thirdly, we use 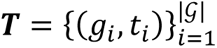 to hold the transcriptomic expression levels of all the genes considered by the model, in which *g*_*i*_ is the name of the *i*th gene and *t*_*i*_ is the quantile-normalized value of its expression level. In this way, the source domain model 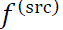 and the target domain model 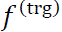 can be obtained by providing one model parameterized by 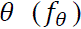 with different condition information, i.e., 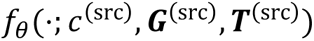 and 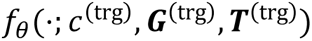, respectively. As the denoising diffusion model is usually implemented as a U-Net architecture ^34^, the two types of condition information are added to each layer of the U-Net model via the categorical embedding and the multi-omics embedding, respectively:

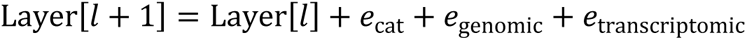

The categorical embeddings are initialized randomly and trained together with the parameters of the U-Net. As for the genomic embeddings, for each gene *g*, His-MMDM utilizes one embedding for the wide-type (WT) version of the gene (*e*_*g*_) and the other for the mutated version of the gene 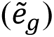 and use a multi-layer perceptron (MLP) to transform them, in other words:

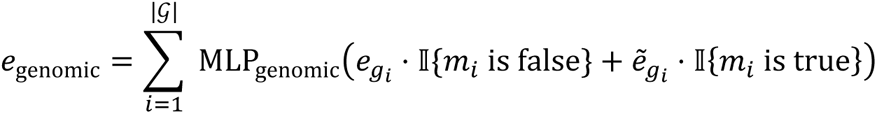

To account for the quantitative effect of transcriptomics, 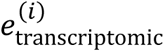 is implemented as two different networks (both as MLPs) that deal with genes and expression levels separately:

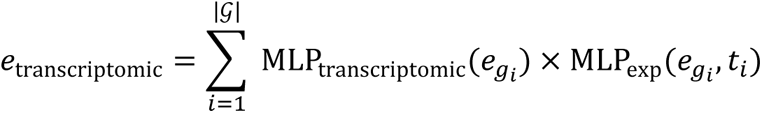

We first pre-train the denoising diffusion model *f*_θ_ using the standard classifier-guided diffusion training procedure on the domains of images between which we wish to translate ^80^. We used the **Algorithm 1** in ^7^ for the training procedure. After *f*_θ_ is successfully trained, we use the denoising diffusion implicit models (DDIMs) as ODESolve for image translation. The process of it can be summarized as **Algorithm 1**. The details of the model architecture, hyperparameters and other configurations regarding training and translation are specified in **Table S1**.

For comparison, we compared the performance of His-MMDM with two GAN-based models, CycleGAN ^28^ and CUT ^37^. To translate images from domain X to domain Y, CycleGAN simultaneously establishes two GANs where one translates images from X to Y and the other from Y to X. To ensure the consistency of the translated images, a cycle-consistency loss is imposed to ensure the invariance when an image is translated from X to Y and then back from Y to X. The other GAN-based model, CUT, resorts to a different approach. CUT establishes only one GAN model to achieve one-way translation from X to Y. It ensures consistency of translation using patchwise contrastive learning ^81,82^ to encourage image patches of the source and translated images of the same location to be similar.

### Image translation tasks and the evaluation of the synthesized images

We summarize the specific settings of image translation tasks, including the specifications of the model checkpoints, the dataset used the condition information of the translation tasks in **Table S1**. The Frechet Inception Distance (FID) ^83^ was used to evaluate the quality of a set of synthesized images against a set of real images. For baseline models (CycleGAN, CUT, D2C, BBDM, and AI-FFPE), we trained and tested them on the same train/test split as His-MMDM. The image features after the last pooling layer of an Inception V3 model ^41^ were used for the computation. Let (μ, Σ) and (μ_0_, Σ_0_) denote the mean and covariance of the Inception V3 features of the synthesized images and real images respectively. The FID score is defined as,

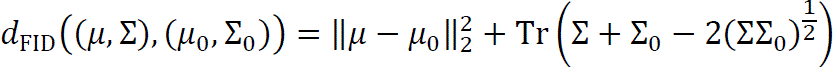

To quantify the effect of image translation from cryosection to FFPE, across primary tumor types, and across tumor organ sites, we computed the difference of FID scores before and after image translation, i.e., 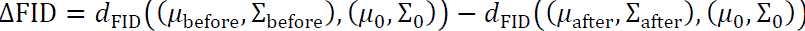. Although both CycleGAN and CUT are GAN-based algorithms, CycleGAN utilizes cycle-consistency for image translation while CUT leverages patch-wise contrastive learning^81,82^. Based on the reported performance on TCGA, although patch-wise contrastive learning is an effective objective to train image translation models between different photography domains, it seems to be less effective in histopathological domains, possibly due to the repetitive nature of the contents of the histopathological images which can confuse this contrastive objective. D2C ^39^ is an image translation model based on latent diffusion. It performs image translation by using a discriminative classifier to guide the sampling process within the latent space. Designed primarily for few-shot translation, D2C keeps the diffusion model, encoder, and decoder fixed during adaptation to new conditions. This makes it well-suited for editing images with distinct, localized regions but less effective for broader image translation tasks, such as those in histopathology (**Fig. 2b**). Similarly, BBDM ^38^ is a diffusion-based model that employs a latent diffusion approach. It utilizes a Brownian bridge in the latent space to enable paired image translation between domains. While BBDM performs comparably to His-MMDM (**Fig. 2b**), its paired formulation makes multi-domain image translation less straightforward.

To compute the magnitude of the effect of genomic– and transcriptomic-guided editing, we used cosine distance rather than FID as we were interested in the editing effect on a single image rather than a collection of images. Besides the cosine distance of the Inception V3 deep features, we also evaluated the difference between image pairs based on their structural similarity index (SSIM) ^84^. SSIM between two image regions *x* and *y* is defined as,

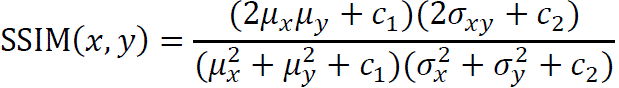

where μ_*x*_ and μ_*y*_ are the pixel mean values of *x* and *y*, 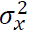 and 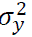 are the pixel variance values of *x* and *y*, σ_*xy*_ is the covariance of *x* and *y*, and *c*_1_ and *c*_2_ are constant parameters. The full SSIM score of two images is computed and averaged using a sliding window approach that fully iterates over the two images, using the implementation from scikit-image ^85^. We convert SSIM into ‘structural distance’ by reversing and scaling it into the range [0,1], using the formula 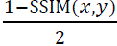. Both the structural distance map (per sliding window) and the structural distance score (averaged for all sliding windows) are used in reporting.

Due to the vast difference between H&E and IHC-stained images, we evaluated the quality of IHC virtual staining in a different way than the previous two tasks. Instead, we computed the ratio of the FID score between two different sets of real IHC images and the FID score between synthesized and real IHC images, i.e., 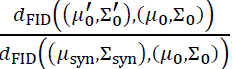. We refer to this as the ‘inverted normalized FID score’ in the main text. In this way, both the effect of image translation and the inherent variability of images of an IHC-stain can be taken into account, and the inversion makes the metric greater if the synthetic IHC images are more similar to the real IHC images.

### Enhancing the performance of pre-trained foundation models in histopathology

Previous studies have demonstrated the practical value of image translation models, which convert cryosectioned slide images into FFPE ones, in enhancing intra-operative cryosectioned slide images ^26,27^. However, in this work, we highlight a different perspective by showing that these image translation models can improve the performance of existing pre-trained foundation models in histopathology. This is especially relevant since many foundation models, such as PLIP ^16^, Prov-GigaPath ^32^, and CHIEF ^33^ have been trained predominantly on vast collections of FFPE slides rather than cryosectioned ones.

Similar to the experiments conducted in PLIP, we performed image-to-image retrieval using image features encoded by the PLIP image encoder. The retrieval process involved searching a database of TCGA FFPE images from 14 sites, based on their cosine distance to the query image. Following the methodology outlined by Chen et al., 2022 ^86^, we assessed retrieval accuracy using a majority voting scheme (“mMV”@k), which measures the accuracy of the predicted tumor site of the query image based on the majority vote of the top-k retrieved images. To evaluate the improvement in PLIP’s performance on text-guided classification using His-MMDM’s translated images on TCGA cryosectioned slides (**Fig. S1a**), we had our pathologists annotate a small subset of image patches from the cryosectioned slides of the TCGA colon cancer cohort, focusing on the five most abundant classes in the Kather colon dataset ^87^. We employed prompts in the format ‘a xx histopathological image,’ where ‘xx’ corresponded to one of the following: ‘adipose,’ ‘background,’ ‘epithelium,’ ‘lymphocyte,’ or ‘mucus.’ The accuracy of the PLIP model was then evaluated based on the consistency between the model’s prediction (the prompt whose PLIP embedding was closest to the image embedding) and our pathologist’s annotations.

In contrast to the multi-modal PLIP model, the CHIEF model ^33^ is an image-only histopathological model pretrained on vast amounts (60,530 WSIs) of real-world data. Although the dataset consists of images from diverse tumor types (19 anatomical sites), they are still mainly FFPE rather than cryosectioned images. We obtained the cryosectioned slide images from the colorectal tumor types (COAD and READ) and the glioma tumor types (GBM and LGG) of TCGA. We then evaluated the fine-tuning performance of microsatellite instability (MSI) and IDH1 mutation status prediction of the pretrained CHIEF model using either the original cryosectioned images or the FFPE images synthesized from them by His-MMDM. The 5-fold cross-validation results are reported.

### Stain quantification

To compute the positivity of the IHC stains in the translated images, we applied the color deconvolution algorithm ^88^ (implemented by the ‘scikit-image’ package ^85^) to the images and obtained the intensity of the two stains components used in our HMU-1st cohort, i.e., hematoxylin (as background) and 3, 3’-diaminobenzidine (DAB, as positive signal). The averaged intensity of the DAB stain is used as the metric of the positivity of the patch.

### Outlier detection of translated histopathological images

When His-MMDM translates an image in the source domain that is unlikely to have a corresponding image in the target domain, the translated image tends to look very eccentric (**Fig. 4c**). To quantify the eccentricity of the translated images, we fitted an outlier detection model (Isolation Forest ^48^, implemented by the ‘scikit-learn’ package ^89^) on the Inception V3 features of them. Using the fitted model, we were able to obtain an ‘inlier score’ in the range of [−1,1] of each sample, where a score close to 1 indicates that the image is a perfect inlier and a score close to –1 indicates that the image is likely to be an outlier. Superimposing this score on the tSNE plot of the Inception V3 features of the translated patch images also shows that the patches with low scores tend to be located on the edge of the tSNE structure. Using the trained His-MMDM model and the fitted outlier detection model, we applied them to the TCGA LUAD and LUSC cohorts. To obtain an inlier score of a WSI, we sampled 50 image patches from it and aggregated their scores by averaging them. Similarly, we obtained the scores of each patient by averaging the inlier scores of all their available WSIs. Finally, we analyzed the relationship between a patient’s inlier score and the patient’s staging and survival information. The Kaplan-Meier method was used to estimate the survival function of the patients under each condition. P values of the fitted Cox proportional hazard models were reported as statistical significance values.

### Additional evaluation of genomics and transcriptomics-guided editing of histopathological images

In addition to qualitatively evaluating genomics– and transcriptomics-guided image editing by comparing and visualizing the edited images against the originals, we implemented supplementary tests to further validate their alignment with real-world data. First, we curated a database of histopathological images, each characterized by a specific somatic mutation or the up/down regulation of transcriptional pathways from MSigDB. These images were sourced from TCGA slides that were reserved exclusively for testing and not used during model training. The selected images represent regions with high epithelial tissue content, as annotated by pathologists. For each somatic mutation or pathway alteration, 200 patch images were stored for each condition based on whole exome sequencing and transcriptomic profiling. This database serves as a comprehensive resource for examining the histopathological appearance of somatic mutations and pathway changes. We then used the original (q) and edited (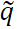) histopathological images as queries to this database. Images ({r_*i*_ })) from a certain condition were retrieved based on the cosine similarity of their PLIP features with the query images. For the top-K retrieved images, we calculated the average cosine distance to assess the overall similarity,

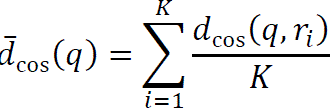

When *q* is the original image without a certain mutation or pathway dysregulation and 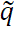 is the His-MMDM-edited image by enabling such conditions, we use the percentage reduction in this metric 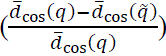 as the effectiveness of image editing. A higher value in this metric indicates better consistency between His-MMDM-edited images to the real images in a particular genomic or transcriptomic condition. *K* is set to 5 in practice.

In addition to the evaluation using the curated database, we further utilized the two sets of images generated by His-MMDM (with or without mutations or pathway up-/down-regulation) as educational examples for pathologists. We randomly sampled real histopathological images from the database, ensuring an equal number of positive and negative classes, and assessed the pathologists’ accuracy in identifying mutations and pathway dysregulations both before and after reviewing the His-MMDM-generated content.

### Statistics

Wilcoxon rank-sum tests and Wilcoxon signed-rank tests were performed using the statistical functions from the Scipy package (scipy.stats) ^90^. Statistical significance (p values) was reported in the respective figures.

**Algorithm 1.**
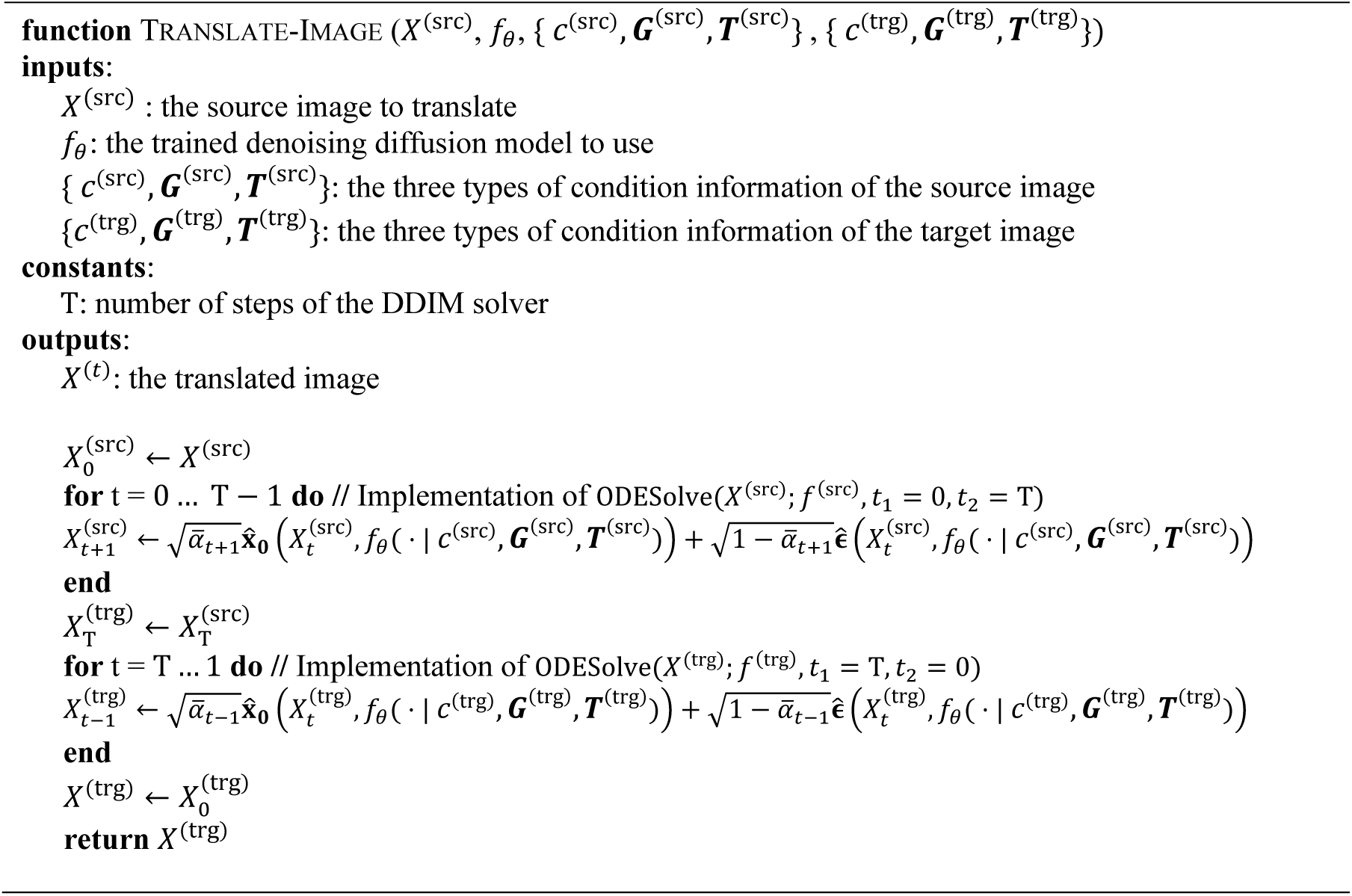
Image-translation with His-MMDM.

## Data Availability

The code and trained checkpoints of His-MMDM are available at https://github.com/lzx325/His-MMDM. Histopathology Data from TCGA used in this study are available from the Genomic Data Commons Portal of the National Cancer Institute (https://gdc.cancer.gov/). Example data from The First Harbin Medical University Hospital and The Harbin Medical University Cancer Hospital are deposited at Zenodo (10.5281/zenodo.12636449) and are available from the lead contact upon reasonable request.

https://zenodo.org/doi/10.5281/zenodo.12636448

## Acknowledgments

This publication is based upon work supported by the King Abdullah University of Science and Technology (KAUST) Office of Research Administration (ORA) under Award No REI/1/5234-01-01, REI/1/5414-01-01, RGC/3/4816-01-01, REI/1/5289-01-01, REI/1/5404-01-01, REI/1/5992-01-01, and URF/1/4663-01-01. The results shown here are in whole or part based upon data generated by the TCGA Research Network: https://www.cancer.gov/tcga.

## Authors’ Contributions

Z.L., T.S., P.L., and X.G. conceived the study. Z.L. and X.G. developed the His-MMDM model and performed the computational analyses, with algorithm development advised by W.H. and computational analysis advised by B.Z. T.S., H.M., S.Z., G.S., Y.W., and P.L. prepared and scanned the histopathological images for the HMU-T dataset, while Y.C., X.C., and S.Z. prepared and scanned the histopathological images for the HMU-1st dataset. Pathologists T.S., Y.C., H.M., and G.S. participated in the inspection and interpretation of the experimental results. Z.L., T.S., and X.G. drafted the majority of the manuscript. All authors reviewed and approved the final manuscript.

---

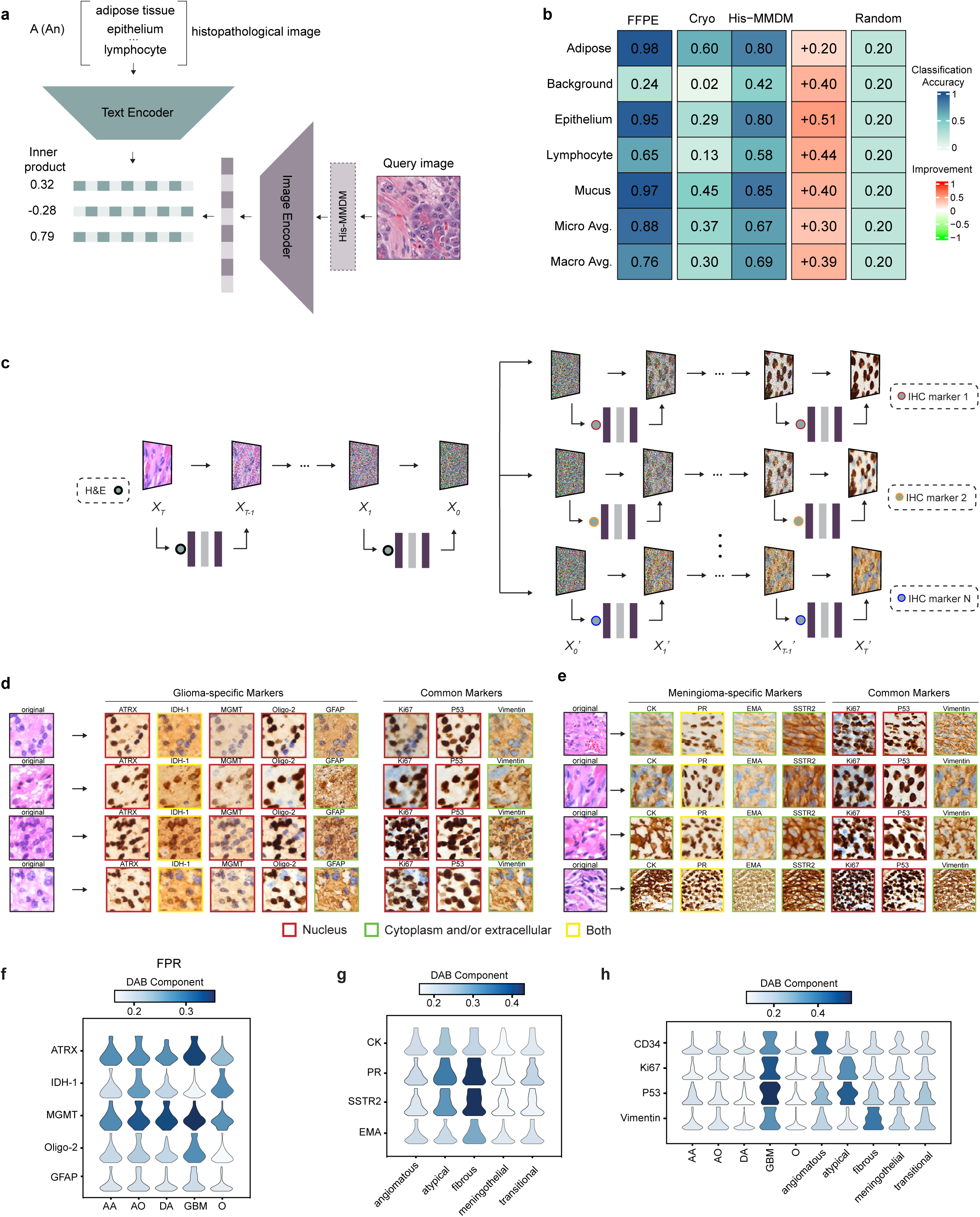

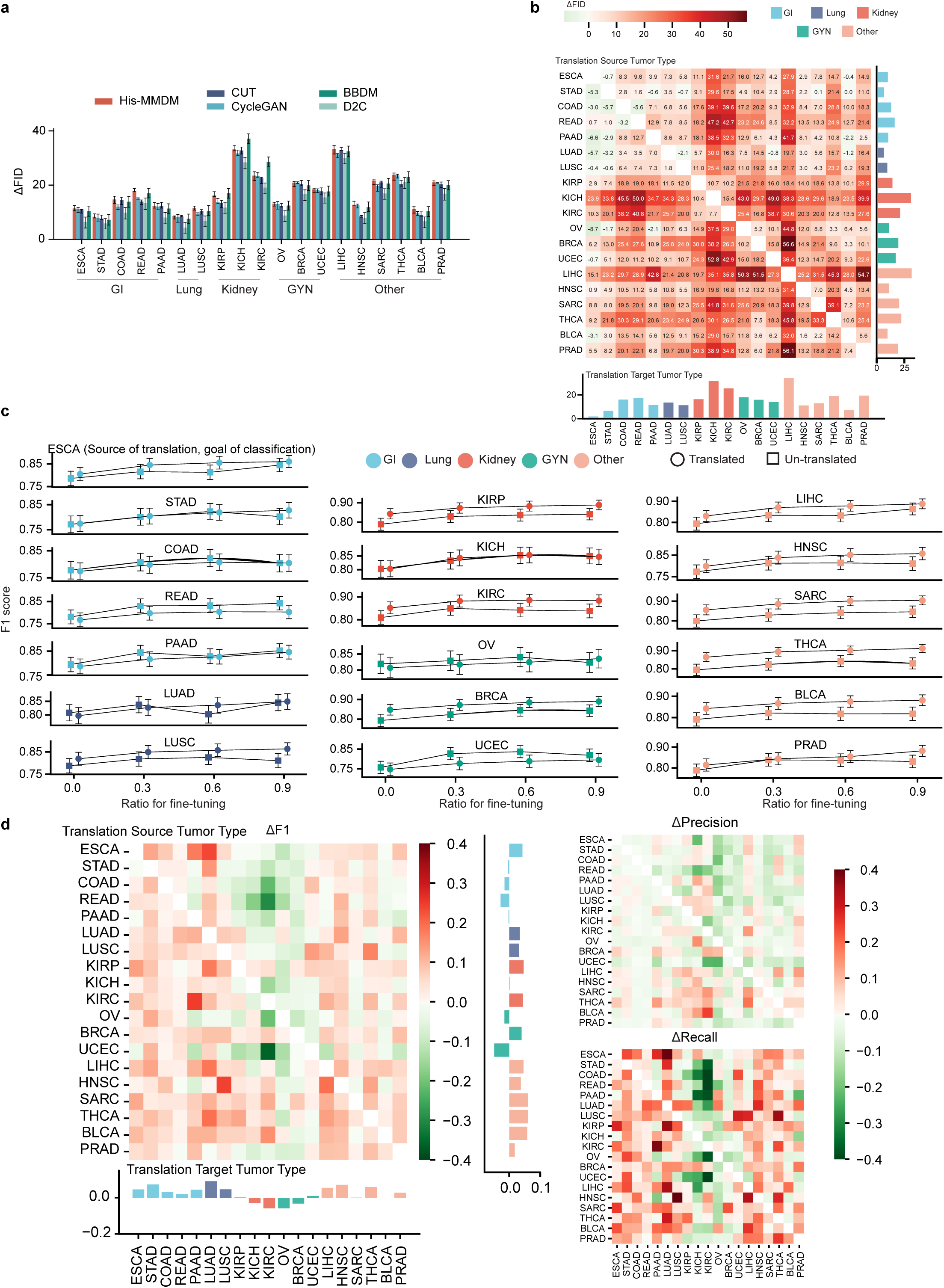

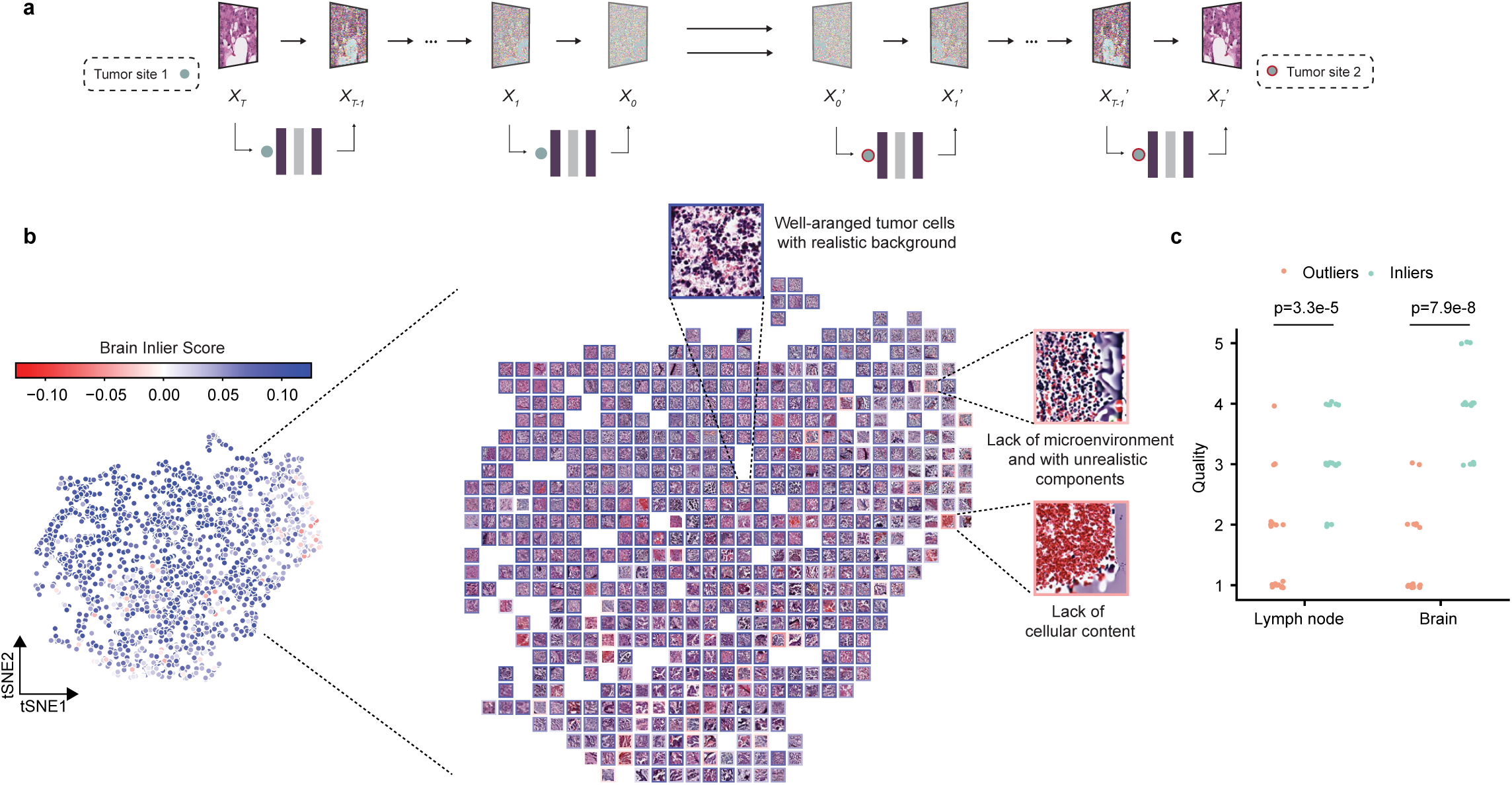

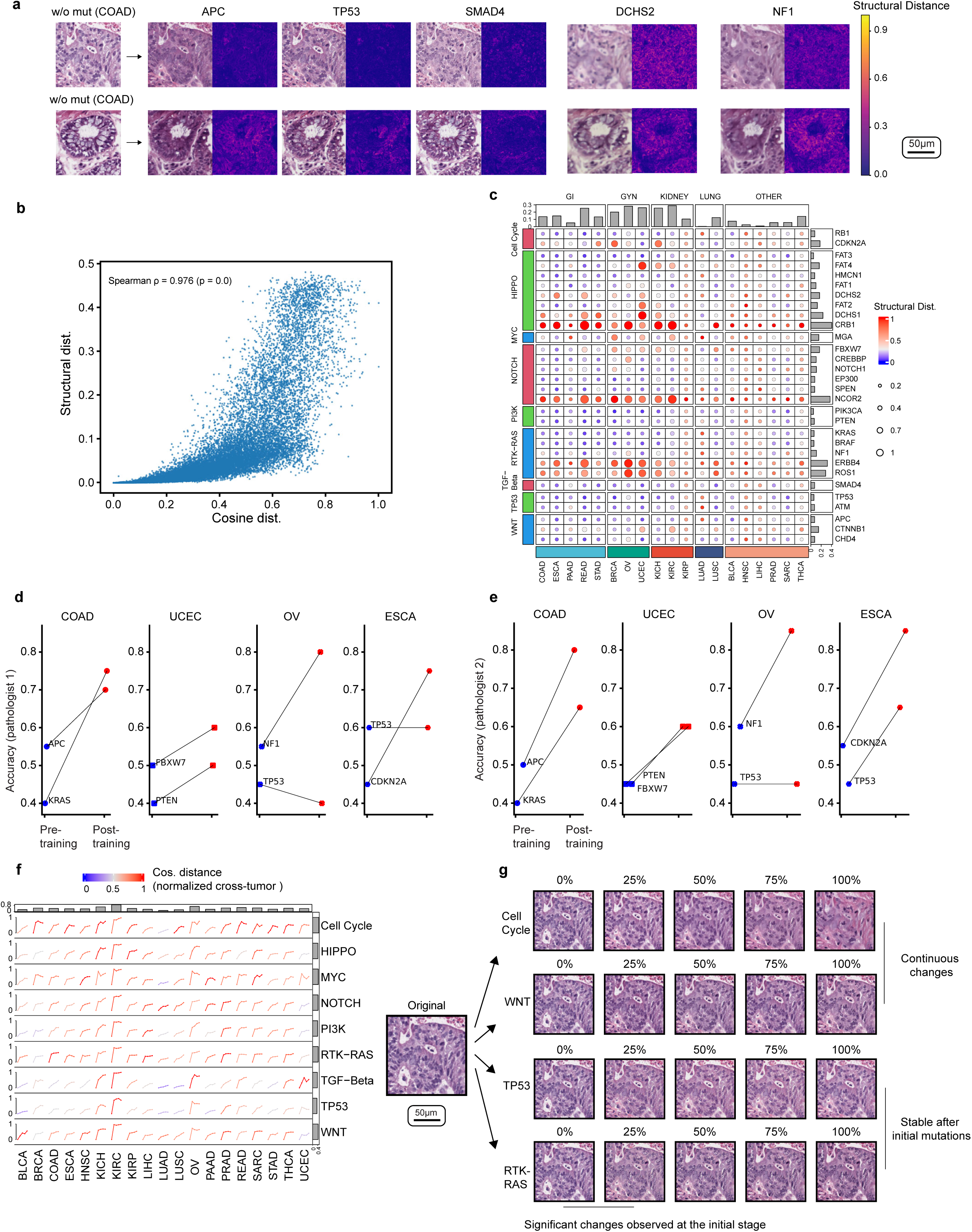

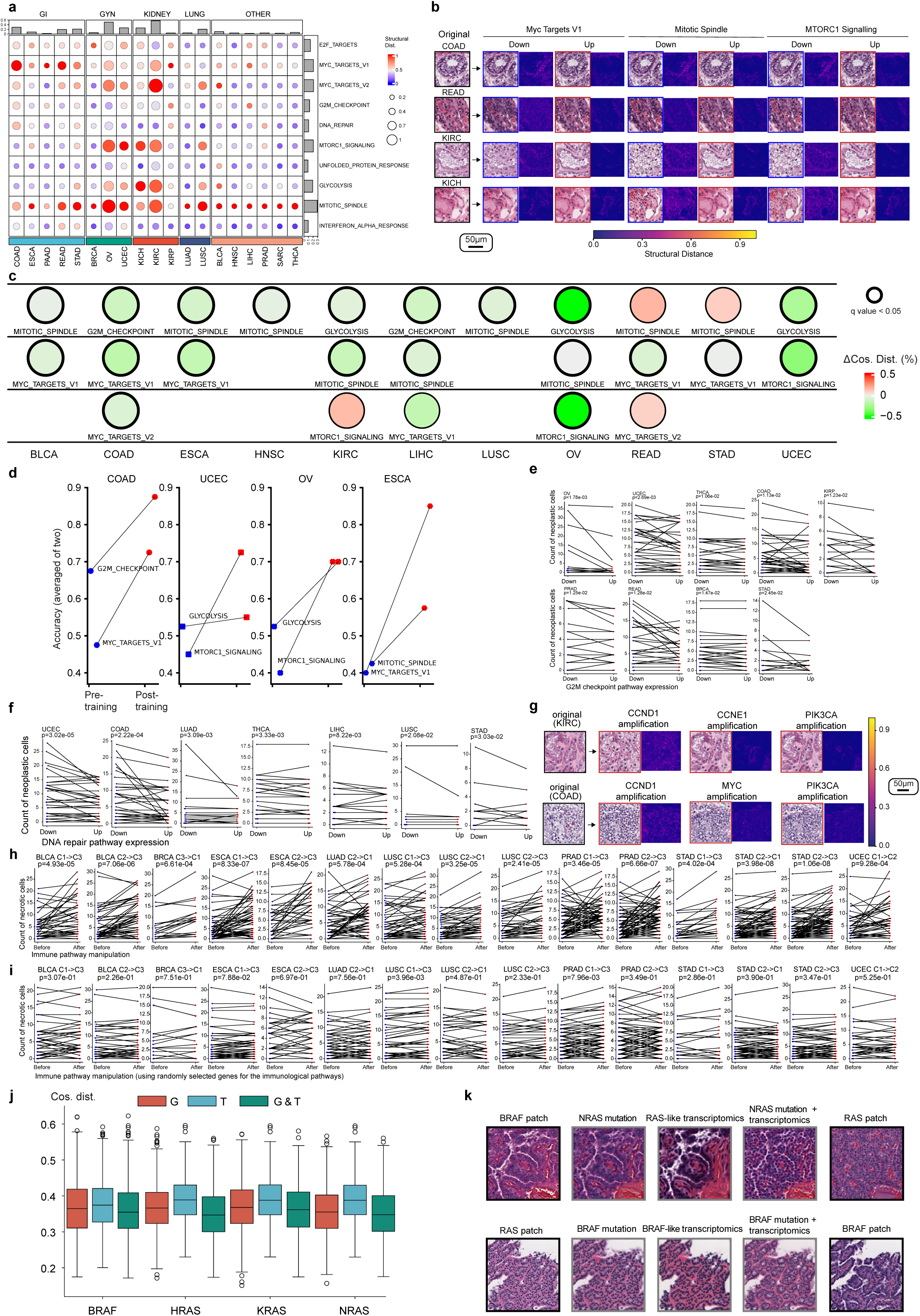

## References

1 Rombach, R., Blattmann, A., Lorenz, D., Esser, P. & Ommer, B. in Proceedings of the IEEE/CVF Conference on Computer Vision and Pattern Recognition. 10684–10695.

2. Midjourney Inc. Midjourney, <www.midjourney.com> (2022).

3 Betker, J. et al. Improving image generation with better captions. Computer Science. https://cdn.openai.com/papers/dall-e-3.pdf2, 8 (2023).

4 Yang, Z. et al. The dawn of lmms: Preliminary explorations with gpt-4v (ision). arXiv preprint arXiv:2309.17421 9, 1 (2023).

5 Zhang, L., Rao, A. & Agrawala, M. in Proceedings of the IEEE/CVF International Conference on Computer Vision. 3836–3847.

6 Goodfellow, I. et al. in Advances in neural information processing systems. 2672–2680.

7 Ho, J., Jain, A. & Abbeel, P. Denoising diffusion probabilistic models. Advances in Neural Information Processing Systems 33, 6840–6851 (2020).

8 Hanna, M. H. et al. Latent transcriptional programs reveal histology-encoded tumor features spanning tissue origins. bioRxiv, 2023.2003.2022.533810, doi:10.1101/2023.03.22.533810 (2023).

9 Moghadam, P. A. et al. in Proceedings of the IEEE/CVF Winter Conference on Applications of Computer Vision. 2000–2009.

10 Shrivastava, A. & Fletcher, P. T. NASDM: Nuclei-Aware Semantic Histopathology Image Generation Using Diffusion Models. arXiv preprint arXiv:2303.11477 (2023).

11 Wong, W. S., Amer, M., Maul, T., Liao, I. Y. & Ahmed, A. in Recent Advances on Soft Computing and Data Mining: Proceedings of the Fourth International Conference on Soft Computing and Data Mining (SCDM 2020), Melaka, Malaysia, January 22– 23, 2020. 392–402 (Springer).

12 Pandey, S., Singh, P. R. & Tian, J. An image augmentation approach using two-stage generative adversarial network for nuclei image segmentation. Biomedical Signal Processing and Control 57, 101782 (2020).

13 Xue, Y. et al. in Medical Image Computing and Computer Assisted Intervention–MICCAI 2019: 22nd International Conference, Shenzhen, China, October 13–17, 2019, Proceedings, Part I 22. 387–396 (Springer).

14 Aversa, M., et al. DiffInfinite: Large Mask-Image Synthesis via Parallel Random Patch Diffusion in Histopathology. arXiv preprint arXiv:2306.13384 (2023).

15. Graikos, A., et al. Learned representation-guided diffusion models for large-image generation. arXiv preprint arXiv:2312.07330 (2023).

16 Huang, Z., Bianchi, F., Yuksekgonul, M., Montine, T. J. & Zou, J. A visual–language foundation model for pathology image analysis using medical Twitter. Nature Medicine, doi:10.1038/s41591-023-02504-3 (2023).

17 Chen, R. J. et al. Towards a general-purpose foundation model for computational pathology. Nature Medicine 30, 850–862, doi:10.1038/s41591-024-02857-3 (2024).

18 Lu, M. Y. et al. A visual-language foundation model for computational pathology. Nature Medicine 30, 863–874, doi:10.1038/s41591-024-02856-4 (2024).

19 Lu, M. Y. et al. A Multimodal Generative AI Copilot for Human Pathology. Nature, doi:10.1038/s41586-024-07618-3 (2024).

20 Zhou, J. et al. Pre-trained multimodal large language model enhances dermatological diagnosis using SkinGPT-4. Nature Communications 15, 5649, doi:10.1038/s41467-024-50043-3 (2024).

21 Zanjani, F. G., Zinger, S., Bejnordi, B. E. & van der Laak, J. A. in 1st Conference on Medical Imaging with Deep Learning (MIDL 2018). 1-11.

22 de Haan, K. et al. Deep learning-based transformation of H&E stained tissues into special stains. Nature Communications 12, 4884, doi:10.1038/s41467-021-25221-2 (2021).

23 Rivenson, Y. et al. Virtual histological staining of unlabelled tissue-autofluorescence images via deep learning. Nature Biomedical Engineering 3, 466–477, doi:10.1038/s41551-019-0362-y (2019).

24 Ghahremani, P. et al. Deep learning-inferred multiplex immunofluorescence for immunohistochemical image quantification. Nature Machine Intelligence 4, 401–412, doi:10.1038/s42256-022-00471-x (2022).

25 Pati, P. et al. Multiplexed tumor profiling with generative AI accelerates histopathology workflows and improves clinical predictions. bioRxiv, 2023.2011.2029.568996, doi:10.1101/2023.11.29.568996 (2023).

26 Ozyoruk, K. B. et al. A deep-learning model for transforming the style of tissue images from cryosectioned to formalin-fixed and paraffin-embedded. Nature Biomedical Engineering 6, 1407–1419 (2022).

27 Liu, Z. et al. Virtual formalin-fixed and paraffin-embedded staining of fresh brain tissue via stimulated Raman CycleGAN model. Science Advances 10, eadn3426, doi:10.1126/sciadv.adn3426 (2024).

28 Zhu, J.-Y., Park, T., Isola, P. & Efros, A. A. in Proceedings of the IEEE international conference on computer vision. 2223–2232.

29 Dhariwal, P. & Nichol, A. Diffusion models beat gans on image synthesis. Advances in neural information processing systems 34, 8780–8794 (2021).

30 Su, X., Song, J., Meng, C. & Ermon, S. Dual diffusion implicit bridges for image-to-image translation. International Conference on Learning Representations (2022).

31 Lu, M. Y., Chen, B. & Mahmood, F. Harnessing medical twitter data for pathology AI. Nature Medicine 29, 2181–2182, doi:10.1038/s41591-023-02530-1 (2023).

32 Xu, H. et al. A whole-slide foundation model for digital pathology from real-world data. Nature, doi:10.1038/s41586-024-07441-w (2024).

33 Wang, X. et al. A pathology foundation model for cancer diagnosis and prognosis prediction. Nature, doi:10.1038/s41586-024-07894-z (2024).

34 Ronneberger, O., Fischer, P. & Brox, T. 234–241 (Springer International Publishing).

35 Song, J., Meng, C. & Ermon, S. (2021).

36. He, B., et al. AI-enabled in silico immunohistochemical characterization for Alzheimer’s disease. Cell Reports Methods 2 (2022).

37 Park, T., Efros, A. A., Zhang, R. & Zhu, J.-Y. in Computer Vision–ECCV 2020: 16th European Conference, Glasgow, UK, August 23–28, 2020, Proceedings, Part IX 16. 319–345 (Springer).

38 Li, B., Xue, K., Liu, B. & Lai, Y.-K. in Proceedings of the IEEE/CVF conference on computer vision and pattern Recognition. 1952–1961.

39 Sinha, A., Song, J., Meng, C. & Ermon, S. D2c: Diffusion-decoding models for few-shot conditional generation. Advances in Neural Information Processing Systems 34, 12533–12548 (2021).

40 Radford, A. et al. in International conference on machine learning. 8748–8763 (PMLR).

41 Szegedy, C., Vanhoucke, V., Ioffe, S., Shlens, J. & Wojna, Z. in Proceedings of the IEEE conference on computer vision and pattern recognition. 2818–2826.

42 Noorbakhsh, J. et al. Deep learning-based cross-classifications reveal conserved spatial behaviors within tumor histological images. Nature Communications 11, 6367, doi:10.1038/s41467-020-20030-5 (2020).

43 Menon, A., Singh, P., Vinod, P. & Jawahar, C. Exploring Histological Similarities Across Cancers From a Deep Learning Perspective. Frontiers in oncology 12 (2022).

44 Quaedvlieg, P. J. F. et al. Histopathological characteristics of metastasizing squamous cell carcinoma of the skin and lips. Histopathology 49, 256–264, 10.1111/j.1365-2559.2006.02472.x (2006).

45 Wang, X. et al. Predicting gastric cancer outcome from resected lymph node histopathology images using deep learning. Nature Communications 12, 1637, doi:10.1038/s41467-021-21674-7 (2021).

46 Huang, S.-C. et al. Deep neural network trained on gigapixel images improves lymph node metastasis detection in clinical settings. Nature Communications 13, 3347, doi:10.1038/s41467-022-30746-1 (2022).

47 Krogue, J. D. et al. Predicting lymph node metastasis from primary tumor histology and clinicopathologic factors in colorectal cancer using deep learning. Communications Medicine 3, 59, doi:10.1038/s43856-023-00282-0 (2023).

48 Liu, F. T., Ting, K. M. & Zhou, Z. H. in 2008 Eighth IEEE International Conference on Data Mining. 413–422.

49 Zou, Y., Bai, H. X., Wang, Z. & Yang, L. Comparison of immunohistochemistry and DNA sequencing for the detection of IDH1 mutations in gliomas. Neuro-oncology 17, 477–478, doi:10.1093/neuonc/nou351 (2015).

50 Bilal, M. et al. Development and validation of a weakly supervised deep learning framework to predict the status of molecular pathways and key mutations in colorectal cancer from routine histology images: a retrospective study. The Lancet Digital Health 3, e763–e772, 10.1016/S2589-7500(21)00180-1 (2021).

51 Chen, M. et al. Classification and mutation prediction based on histopathology H&E images in liver cancer using deep learning. npj Precision Oncology 4, 14, doi:10.1038/s41698-020-0120-3 (2020).

52 Jain, M. S. & Massoud, T. F. Predicting tumour mutational burden from histopathological images using multiscale deep learning. Nature Machine Intelligence 2, 356–362, doi:10.1038/s42256-020-0190-5 (2020).

53 Coudray, N. et al. Classification and mutation prediction from non–small cell lung cancer histopathology images using deep learning. Nature Medicine 24, 1559–1567, doi:10.1038/s41591-018-0177-5 (2018).

54 Qu, H. et al. Genetic mutation and biological pathway prediction based on whole slide images in breast carcinoma using deep learning. NPJ Precis Oncol 5, 87, doi:10.1038/s41698-021-00225-9 (2021).

55 Li, Z. et al. Vision transformer-based weakly supervised histopathological image analysis of primary brain tumors. iScience 26, 105872, doi:10.1016/j.isci.2022.105872 (2023).

56 Fu, Y. et al. Pan-cancer computational histopathology reveals mutations, tumor composition and prognosis. Nature Cancer 1, 800–810, doi:10.1038/s43018-020-0085-8 (2020).

57 Schmauch, B. et al. A deep learning model to predict RNA-Seq expression of tumours from whole slide images. Nature communications 11, 1–15 (2020).

58 Sanchez-Vega, F. et al. Oncogenic Signaling Pathways in The Cancer Genome Atlas. Cell 173, 321–337.e310, 10.1016/j.cell.2018.03.035 (2018).

59 Subramanian, A. et al. Gene set enrichment analysis: a knowledge-based approach for interpreting genome-wide expression profiles. Proceedings of the National Academy of Sciences 102, 15545–15550 (2005).

60 Liberzon, A. et al. The Molecular Signatures Database Hallmark Gene Set Collection. Cell Systems 1, 417–425, 10.1016/j.cels.2015.12.004 (2015).

61 Graham, S. et al. Hover-Net: Simultaneous segmentation and classification of nuclei in multi-tissue histology images. Medical Image Analysis 58, 101563, 10.1016/j.media.2019.101563 (2019).

62 Cai, H. et al. CRISPR/Cas9 model of prostate cancer identifies Kmt2c deficiency as a metastatic driver by Odam/Cabs1 gene cluster expression. Nature Communications 15, 2088, doi:10.1038/s41467-024-46370-0 (2024).

63 Aaltonen, L. A. et al. Pan-cancer analysis of whole genomes. Nature 578, 82–93, doi:10.1038/s41586-020-1969-6 (2020).

64 Thorsson, V. et al. The Immune Landscape of Cancer. Immunity 48, 812–830.e814, 10.1016/j.immuni.2018.03.023 (2018).

65 Lu, M. Y. et al. AI-based pathology predicts origins for cancers of unknown primary. Nature 594, 106–110, doi:10.1038/s41586-021-03512-4 (2021).

66 Dolezal, J. M. et al. Deep learning generates synthetic cancer histology for explainability and education. npj Precision Oncology 7, 49, doi:10.1038/s41698-023-00399-4 (2023).

67 Carrillo-Perez, F. et al. Synthetic whole-slide image tile generation with gene expression profile-infused deep generative models. Cell Reports Methods 3, doi:10.1016/j.crmeth.2023.100534 (2023).

68 Carrillo-Perez, F. et al. Generation of synthetic whole-slide image tiles of tumours from RNA-sequencing data via cascaded diffusion models. Nature Biomedical Engineering, doi:10.1038/s41551-024-01193-8 (2024).

69. AutoGPT, <https://news.agpt.co/> (2023).

70 Cao, Y., et al. A comprehensive survey of ai-generated content (aigc): A history of generative ai from gan to chatgpt. arXiv preprint arXiv:2303.04226 (2023).

71 Roziere, B. et al. Code llama: Open foundation models for code. arXiv preprint arXiv:2308.12950 (2023).

72. Lu, M. Y., et al. A Foundational Multimodal Vision Language AI Assistant for Human Pathology. arXiv preprint arXiv:2312.07814 (2023).

73 Zhou, J. et al. An AI Agent for Fully Automated Multi-omic Analyses. bioRxiv, 2023.2009. 2008.556814 (2023).

74 Ulhaq, A., Akhtar, N. & Pogrebna, G. Efficient diffusion models for vision: A survey. arXiv preprint arXiv:2210.09292 (2022).

75 Lu, C. et al. Dpm-solver: A fast ode solver for diffusion probabilistic model sampling in around 10 steps. Advances in Neural Information Processing Systems 35, 5775–5787 (2022).

76 Lu, C., et al. Dpm-solver++: Fast solver for guided sampling of diffusion probabilistic models. arXiv preprint arXiv:2211.01095 (2022).

77 Goldman, M. et al. The UCSC Xena platform for public and private cancer genomics data visualization and interpretation. biorxiv, 326470 (2018).

78 Krizhevsky, A., Sutskever, I. & Hinton, G. E. in Advances in neural information processing systems. 1097–1105.

79 Dosovitskiy, A., et al. An Image is Worth 16×16 Words: Transformers for Image Recognition at Scale. CoRR abs/2010.11929 (2020).

80 Nichol, A. Q. & Dhariwal, P. 8162–8171 (2021).

81 Henaff, O. in International conference on machine learning. 4182–4192 (PMLR).

82 Bachman, P., Hjelm, R. D. & Buchwalter, W. Learning representations by maximizing mutual information across views. Advances in neural information processing systems 32 (2019).

83 Heusel, M., Ramsauer, H., Unterthiner, T., Nessler, B. & Hochreiter, S. Gans trained by a two time-scale update rule converge to a local nash equilibrium. Advances in neural information processing systems 30 (2017).

84 Wang, Z., Bovik, A. C., Sheikh, H. R. & Simoncelli, E. P. Image quality assessment: from error visibility to structural similarity. IEEE transactions on image processing 13, 600–612 (2004).

85 Van der Walt, S. et al. scikit-image: image processing in Python. PeerJ 2, e453 (2014).

86. Chen, C., et al. Fast and Scalable Image Search For Histology. arXiv preprint arXiv:2107.13587 (2021).

87 Kather, J. N. et al. Predicting survival from colorectal cancer histology slides using deep learning: A retrospective multicenter study. PLOS Medicine 16, e1002730, doi:10.1371/journal.pmed.1002730 (2019).

88 Ruifrok, A. C. & Johnston, D. A. Quantification of histochemical staining by color deconvolution. Analytical and quantitative cytology and histology 23, 291–299 (2001).

89 Pedregosa, F. et al. Scikit-learn: Machine learning in Python. the Journal of machine Learning research 12, 2825–2830 (2011).

90 Virtanen, P. et al. SciPy 1.0: fundamental algorithms for scientific computing in Python. Nature methods 17, 261–272 (2020).

