## Supplementary Notes and Legends for "His-MMDM: Multi-domain and Multi-omics Translation of Histopathological Images with Diffusion Models"

#### Section S1 Additional results of IHC virtual staining

Using color deconvolution, we computed the intensity of the DAB stain on the virtually stained images in each glioma and meningioma subtype (**Fig. S1f-S1h**). Most notably, the positivity of IDH-1 (suggesting IDH-1 mutation) is higher in lower-grade glioma subtypes (AA, AO, DA, and O) rather than in glioblastomas (GBM), which is consistent with previous findings that IDH-1 mutations are more frequently found in the lower-grade gliomas or secondary GBMs that stem from them, rather than the primary GBMs [1] (**Fig. S1f**). The intensity of subtype-nonspecific markers tends to correlate with the proliferation activity and cell density of the specific subtypes. For example, Oligo-2, as a transcription factor necessary in the development of nearly all glioma subtypes [2], tends to have a higher intensity in the more proliferative GBM subtype (**Fig. S1f**). Similarly, EMA[3], PR [4], and SSTR2 [5], being common meningioma markers, tend to have higher intensity in the more proliferative atypical or the more dense fibrous subtype (**Fig. S1g**). As for the common markers used in both glioma and meningioma, CD34, an endothelium marker, reasonably exhibits a higher intensity in angiomatous meningioma and in GBM where angiogenesis is frequent (**Fig. S1k**) [6]. Additionally, GBM and the higher-grade atypical meningioma also show higher intensity of the cell proliferation marker Ki67 (suggesting higher expression) and tumor suppressor p53 (suggesting p53 mutation). Collectively, these analyses demonstrate the effectiveness of His-MMDM in virtual staining and the consistency of the synthesized images with prior knowledge.

#### Section S2 Additional results of primary tumor type translation

As a comparison, we also trained two GAN-based models, CycleGAN [7] and CUT [8] (**Methods**), and two other diffusion model-based methods D2C [9] and BBDM [10] (**Methods**). The Frechet Inception Distance (FID) [11] of the translated images to a particular tumor type and the real images of that tumor type were computed as the evaluation of image fidelity (the *lower* FID scores imply *higher* fidelity) (**Methods**). We further computed the FID scores of the real un-translated images between tumor types and computed the reduction of this metric ($\Delta\mathrm{FID} = \mathrm{FID}_{\mathrm{before}} - \mathrm{FID}_{\mathrm{after}}$) before and after translation by His-MMDM. Overall, His-MMDM achieved similar or even better performance (ΔFID) compared to its GAN-based counterparts, especially for tumor types such as BLCA, KIRP, and LUSC (**Fig. S2a**). His-MMDM achieved comparable performance as BBDM and is better than D2C. The largest ΔFIDs are observed between very distant tumor types, such as between the GYN tumors and kidney tumors (**Fig. S2b**). In particular, due to the distinctiveness of kidney and liver tumors, their ΔFID values are the highest when other tumors are either translated to or from them (**Fig. S2b**). Previous cross-classification studies using discriminative models reported high performance between related tumor types, such as within the GI tumors or the lung tumors [12]. Consistently, we also observed that the ΔFID values tend to be the lowest within the GI tumors and relatively low within the GYN tumors, kidney, and lung tumors.

We compared the performance (in terms of F1 score) of binary tumor classification models trained on one tumor type (the target of translation) and cross-classifying images from another tumor type (the source of translation/the goal of classification) using either the original images or the translated images. The classification performance in four out of five tumor families corresponding to 12 out of 19 tumor types on average showed higher performance using the translated images (**Fig. 3c** and **Fig. S2c**, 'ratio for fine-tuning = 0'). Additionally, to improve the classification models' performance, we fine-tuned the classification models using 30%, 60%, and 90% of the translated images (**Fig. 3c** and **Fig. S2c**, 'ratio for fine-tuning = 0.3, 0.6, 0.9'). The number of tumor types with higher performance using the translated images increases to 13 at 60% and 15 at 90%. The performance improvement is most evident when translating from the GI tumors to the lung tumors, and the improvement is more in recall than in precision (**Fig. S2d**). This improvement in recall could be due to the adaptation of the features of the unknown-type image into known ones. Therefore, translating images from unseen tumor types to known tumor types is indeed beneficial for a discriminative model's inference and further fine-tuning.

**Section S3 Editing between BRAF-like and RAS-like thyroid tumors**

We finally demonstrate through an example when His-MMDM is guided by the combination of genomic mutations and transcriptomic profiles to edit histopathological images. In thyroid tumors, the *BRAF*-*RAS* score is used to distinguish classical thyroid tumors characterized by the $BRAF^{V600E}$ mutation (*BRAF*-like) from the less malignant follicular variant that harbors *RAS* mutations (*RAS*-like). Very recently, a GAN-based generative model has been developed by Dolezal et al. to generate thyroid histopathological images across the full spectrum of the *BRAF*-*RAS* score (ranging from -1 (the most *BRAF*-like) to 1 (the most *RAS*-like)) [13]. One highlight of Dolezal et al.'s model is that once a particular random seed is set, the model can produce *BRAF*-like and *RAS*-like images *that are paired* (see Fig. 3 of Dolezal et al. [13]). We, therefore, investigated whether His-MMDM can recapitulate the patterns of *BRAF*-like and *RAS*-like thyroid tumors learned by the Dolezal et al. model. We first generated paired BRAF-like (score = -1) and RAS-like (score = 1) images using the Dolezal et al. model. Then we used His-MMDM to edit the *BRAF*-like images into *RAS*-like ones and the *RAS*-like images into *BRAF*-like ones, by modifying their genomic mutations (in either *BRAF*, *NRAS*, *HRAS* or *KRAS*) or manipulating their transcriptomic profiles (according to their respective transcriptomic signatures) or both (**Methods**). By comparing the His-MMDM-edited images to the other 'ground truth' image in the pair generated from the Dolezal et al. model, we were able to dissect individual contributions from the genomic side or the transcriptomic side, and the combined genomic and transcriptomic guidance resulted in higher similarity of the His-MMDM edited images to the 'ground truth' images (**Fig. S5j**). By visualizing the edited images, it is evident that His-MMDM recapitulated the arrangement and the characteristic structural features of thyroid tumors of each subtype, e.g., the papillary structures of the *BRAF*-like and the follicular structures of the *RAS*-like) (**Fig. S5k**).

### Supplementary Figures

**Figure S1 Cryosection to FFPE conversion and virtual IHC staining of histopathological images. Related to Figure 2.**

(a) The schematic illustrating the usage of His-MMDM converted cryosectioned slide images for text-guided classification using the pre-trained multi-modal foundation model, PLIP.

(b) His-MMDM-converted cryosectioned slide images improve the text-guided classification accuracy of the PLIP model. FFPE: using FFPE images, Cryo: using original cryosection images, His-MMDM: using His-MMDM translated images

(c) A schematic that illustrates how His-MMDM achieves virtual staining of multiple IHC markers.

(d) More examples of virtual staining of glioma images, by both the glioma-specific IHC markers and the common IHC markers.

(e) More examples of virtual staining of meningioma images, by both the glioma-specific IHC markers and the common IHC markers.

(f-h) The violin plots showing the intensity of the positive (DAB) stain on the virtually-stained images of glioma-specific markers (f) meningioma-specific markers (g) and common markers (h) among the primary brain tumor subtypes.

**Figure S2 Translation of histopathological images across primary tumor types. Related to Figure 3.**

(a) Evaluation of His-MMDM (in terms of ΔFID scores) in performing cross-tumor type translation and comparison with other image translation models.

(b) Detailed pairwise heatmap showing the improvement of FID score ($\Delta\text{FID} = \text{FID}_{\text{before}} - \text{FID}_{\text{after}}$) between each pair of TCGA classes. Bar plots show the averaged $\Delta\text{FID}$ for a particular translation source tumor type (rows) or target tumor type (columns).

(c) The classification performance (F1 score) of binary tumor classifiers that are trained on other tumor types (as targets of translation) using either translated/untranslated images when there is no additional fine-tuning ('Ratio for fine-tuning = 0') or with additional fine-tuning ('ratio for fine-tuning = 0.3, 0.6, 0.9') in each tumor type (as the source of translation and the goal of classification). Data for each source of translation is displayed separately.

(d) Improvement of classification performance (F1 score, precision, and recall) of binary tumor classifiers in one tumor type (as the source of translation, rows) that are trained on other tumor types (as the targets of translation, columns). Improvement is computed as the improvement of the metrics after image translation by His-MMDM.

**Figure S3 Translation of histopathological images between primary and metastatic organ sites. Related to Figure 4.**
(a) A schematic that illustrates how His-MMDM achieves histopathological image translation between different primary organ sites.

(b) Visualization of running the outlier detection model (Isolation Forest) on the translated images from the lung to the brain. The images are visualized according to their structure in the tSNE space and their 'inlier scores' are superimposed.

(c) Pathologist-assessed visual quality on randomly selected translated images. For the images translated to lymph node and brain, 20 images were randomly selected from the inliers (in the interior of the tSNE grid) and outliers (on the edge of the tSNE grid), and a pathologist ranked the visual quality of each of them (on a 1-5 scale).

**Figure S4 The genomics-guided editing of histopathological images. Related to Figure 5.**
(a) Example COAD images edited into different single genomic mutations (*APC*, *TP53*, *SMAD4*, *DCHS2*, and *NF1*). Structural distance maps are shown to highlight the differences.

(b) The structural distance and cosine distance (InceptionV3 feature) between edited images have a very high correlation (Spearman $\rho = 0.976$).

(c) The effect of genomic mutations on histopathological images by each tumor type, measured by the structural distance between the WT version image and the mutated image.

(d) Performance of the first pathologist's recognition of the top genomic mutations in four tumors (COAD, UCEC, OV, and ESCA) before and after observing His-MMDM's generated images as a tutorial. Improvement is observed except for TP53 in OV.

(e) Performance of the second pathologist's recognition of the top genomic mutations in four tumors (COAD, UCEC, OV, and ESCA) before and after observing His-MMDM's generated images as a tutorial. Improvement is observed except for TP53 in OV.

(f) Cosine distances of images edited with sequentially accumulating mutations to the WT version of the image. The images sequentially (sorted by mutation rate from high to low) performed the mutation of 25%, 50%, 75%, and 100% of genes in each oncological pathway. The cosine distances were computed w.r.t. the WT version (0% mutation) and visualized through each line plot. Each line plot is colored with the median cosine distance (cross-tumor normalized) when 25%, 50%, 75%, and 100% were mutated. Bar plots aggregate per each pathway (row) or tumor type (row).

(g) Example COAD images edited into accumulating genomic mutations (0%, 25%, 50%, 75%, and 100% of genes, ordered by mutation rate) of four different oncological pathways (Cell Cycle, WNT, TP53, and RTK-RAS).

**Figure S5 The transcriptomics-guided editing of histopathological images. Related to Figure 6.**

(a) The effect of transcriptional pathway manipulations on histopathological images by each tumor type, measured by the structural distance between the WT version image and mutated image.

(b) Performance of pathologists' recognition of the top pathway alterations in four tumors (COAD, UCEC, OV, and ESCA) before and after observing His-MMDM's generated images as a tutorial.

(c) The computation of the change in cosine distance of a query image with a certain pathway down-regulated to its nearest neighbors in the database when the image is edited into a version with that pathway is up-regulated. 20 out of the top 25 tumor type and pathway pairs where His-MMDM's edits resulted in the most notable feature changes displayed a statistically significant reduction in the distance to their nearest neighbors. q values are from the Wilcoxon signed-rank test adjusted by the Benjamini–Hochberg procedure.

(d) Examples of MSigDB transcriptomic pathway manipulations.

(e) The complete tumor types in which the knock-up of the pathway ‘G2M checkpoint’ reduces the number of malignant cells detected by Hover-Net.

(f) The complete tumor types in which knock-up of the pathway ‘DNA repair’ reduces the number of malignant cells detected by Hover-Net.

(g) Examples of a KIRC image and a COAD image edited into somatic copy number amplification of different genes.

(h) The complete list of translations between immune subtypes that increase the count of necrotic cells detected by Hover-Net. Each dot represents a particular image before (blue) or after (red) translation. P values from the Wilcoxon signed rank test.

(i) The same set of translations between the immune subtypes and in the tumor types as (f), but performed with randomly sampled genes. Each dot represents a particular image before (blue) or after (red) translation. Almost all translations showed insignificant changes in the count of necrotic cells before and after translation (except LUSC C1→C3 and PRAD C2→C3 showed some levels of significance). P values from the Wilcoxon signed rank test.

(j) Translation of THCA images from BRAF-like to RAS-like and vice versa. Cosine distances of the edited THCA images using either genomics alone (G), transcriptomics alone (T), or both (GT) to the ‘ground truth’ images of Dolezal et al.’s model are reported. When translating in the direction from BRAF-like to RAS-like, there are three options for genomic mutations (HRAS, KRAS, and NRAS), compared to the only option (BRAF) in the opposite direction.

(k) Histopathological image examples illustrating the effect of editing images from BRAF-like to RAS-like and vice versa using either genomics alone, transcriptomics alone, or both.

### Supplementary Tables

**Table S1 The model architecture, hyperparameters, training configurations, and task specifications.**
(a) The detailed architecture of the U-Net model in His-MMDM

(b) The hyperparameters used for training and testing

(c) Summary of the image translation tasks for His-MMDM

**Table S2 Dataset statistics.**
(a-c) Statistics of the TCGA dataset (a), the HMU-C dataset (b), and the HMU-1st dataset (c)

**Table S3 Abbreviations of tumor types.**

**Table S4 Description of the IHC markers used in virtual staining.**

**Table S5 Relevant genes and pathways used in the experiments.**
(a) The genes used in the genomic embeddings of His-MMDM. Each gene is marked for (1) its membership in oncological pathways defined in Sanchez-Vega et al. [14] (2) the TCGA cohorts in which they have high mutation rates (3) high discriminative performance reported by Fu et al. [15]. The genes are sorted according to their pan-cancer mutation rate in TCGA.

(b) The genes used in the transcriptomic embeddings of His-MMDM. Each gene is marked for (1) its membership in the MSigDB pathways [16,17] (2) whether they are highly dysregulated in TCGA tumor samples (t-statistic) (3) its membership in the well-predicted gene signatures of HE2RNA [18].

(c) The component genes of the MSigDB pathways.

(d) The definition of the immune subtypes (C1, C2, and C3) in terms of their expression signatures of five immunological pathways.

(e) The component genes of the immunological pathways.

(f) The randomly selected genes for the immunological pathways as a negative control.

(g) The genes with mutations (genomics) and the genes up-regulated (transcriptomics) in the *BRAF*-like and *RAS*-like thyroid tumors.

### References

1 Balss, J. *et al.* Analysis of the IDH1 codon 132 mutation in brain tumors. *Acta Neuropathol* **116**, 597-602, doi:10.1007/s00401-008-0455-2 (2008).
2 Tsigelny, I. F., Kouznetsova, V. L., Lian, N. & Kesari, S. Molecular mechanisms of OLIG2 transcription factor in brain cancer. *Oncotarget* **7**, 53074-53101, doi:10.18632/oncotarget.10628 (2016).
3 Petricevic, J. *et al.* Expression of nestin, mesothelin and epithelial membrane antigen (EMA) in developing and adult human meninges and meningiomas. *Acta histochemica* **113**, 703-711 (2011).
4 Maiuri, F. *et al.* Progesterone Receptor Expression in Meningiomas: Pathological and Prognostic Implications. *Front Oncol* **11**, 611218, doi:10.3389/fonc.2021.611218 (2021).
5 Wu, W. *et al.* Clinical Significance of Somatostatin Receptor (SSTR) 2 in Meningioma. *Front Oncol* **10**, 1633, doi:10.3389/fonc.2020.01633 (2020).
6 Ahir, B. K., Engelhard, H. H. & Lakka, S. S. Tumor Development and Angiogenesis in Adult Brain Tumor: Glioblastoma. *Mol Neurobiol* **57**, 2461-2478, doi:10.1007/s12035-020-01892-8 (2020).
7 Zhu, J.-Y., Park, T., Isola, P. & Efros, A. A. in *Proceedings of the IEEE international conference on computer vision.* 2223-2232.
8 Park, T., Efros, A. A., Zhang, R. & Zhu, J.-Y. in *Computer Vision–ECCV 2020: 16th European Conference, Glasgow, UK, August 23–28, 2020, Proceedings, Part IX 16.* 319-345 (Springer).
9 Sinha, A., Song, J., Meng, C. & Ermon, S. D2c: Diffusion-decoding models for few-shot conditional generation. *Advances in Neural Information Processing Systems* **34**, 12533-12548 (2021).
10 Li, B., Xue, K., Liu, B. & Lai, Y.-K. in *Proceedings of the IEEE/CVF conference on computer vision and pattern Recognition.* 1952-1961.
11 Heusel, M., Ramsauer, H., Unterthiner, T., Nessler, B. & Hochreiter, S. Gans trained by a two time-scale update rule converge to a local nash equilibrium. *Advances in neural information processing systems* **30** (2017).
12 Noorbakhsh, J. *et al.* Deep learning-based cross-classifications reveal conserved spatial behaviors within tumor histological images. *Nature Communications* **11**, 6367, doi:10.1038/s41467-020-20030-5 (2020).
13 Dolezal, J. M. *et al.* Deep learning generates synthetic cancer histology for explainability and education. *npj Precision Oncology* **7**, 49, doi:10.1038/s41698-023-00399-4 (2023).
14 Sanchez-Vega, F. *et al.* Oncogenic Signaling Pathways in The Cancer Genome Atlas. *Cell* **173**, 321-337.e310, doi:https://doi.org/10.1016/j.cell.2018.03.035 (2018).
15 Fu, Y. *et al.* Pan-cancer computational histopathology reveals mutations, tumor composition and prognosis. *Nature Cancer* **1**, 800-810, doi:10.1038/s43018-020-0085-8 (2020).
16 Subramanian, A. *et al.* Gene set enrichment analysis: a knowledge-based approach for interpreting genome-wide expression profiles. *Proceedings of the National Academy of Sciences* **102**, 15545-15550 (2005).
17 Liberzon, A. *et al.* The Molecular Signatures Database Hallmark Gene Set Collection. *Cell Systems* **1**, 417-425, doi:https://doi.org/10.1016/j.cels.2015.12.004 (2015).
18 Schmauch, B. *et al.* A deep learning model to predict RNA-Seq expression of tumours from whole slide images. *Nature communications* **11**, 1-15 (2020).
